# Childhood immune imprinting shapes cohort and period influenza mortality

**DOI:** 10.64898/2025.12.03.25341584

**Authors:** Kylee A. Hoffman, Chadi M. Saad-Roy, Ayesha S. Mahmud

## Abstract

Influenza viruses encountered in childhood can leave a lasting immunological imprint. To disentangle the mortality effects of age, the circulating seasonal strain, and immune history, we fit statistical mortality models to 54 years of influenza mortality data from the U.S. We find strong signatures of subtype-level imprinting in H1N1-dominated seasons following the 2009 pandemic – cohorts imprinted with more similar H1N1 strains experience greater protection. Furthermore, we find large differences in age-specific mortality risk across cohorts based on their imprinted strain and the seasonal strains that were dominant throughout their lifetime. In contrast to older H1N1- and H2N2-imprinted cohorts, our results show that more recent cohorts imprinted with H3N2 have experienced substantially higher mortality through most of their lifetime. Our results highlight the long-term consequences of immune imprinting, and its impact on period and cohort influenza mortality. Overall, our findings have important implications for vaccination efforts and future influenza mortality burden.

---

Since the early twentieth century, influenza A virus (IAV) pandemics and seasonal outbreaks have shaped observable variations in exposure and immune memory across human populations, influencing cohort-specific susceptibility to different IAV strains. Two glycoproteins on the surface of influenza viruses – hemagglutinin (HA) and neuraminidase (NA) – contain the major antigenic sites targeted by antibodies and are the key determinants of susceptibility to infection. The process of antigenic drift, i.e. the accumulation of HA and NA mutations from one season to the next, allows influenza viruses to evade hosts’ immunity and can cause repeat infections throughout an individual’s lifetime. Antigenic shift, i.e. the reassortment of HA and NA proteins, can introduce a novel IAV into human populations and cause pandemics.

There is a growing body of evidence showing that influenza viruses encountered in childhood can leave a lasting immunological imprint, with lifelong impacts on an individual’s adaptive immune response to influenza infections (*1–6*). Seminal work on the “original antigenic sin” showed that individuals have the highest antibody titers against the influenza strains first encountered in childhood (*1*). More recent work has shown that childhood immune imprinting protects against novel avian influenza viruses of the same phylogenetic HA group as the first childhood exposure, and against seasonal influenza viruses of the same subtype as the first childhood exposure (*2–4, 6–9*). Other recent studies have found evidence for antigenic seniority, in which the immune system continually enhances antibody levels against the strains encountered in childhood more than against strains encountered later in life (*10, 11*).

The recorded history of IAV circulation in human populations can be characterized into several distinct phases, each dominated by a particular phylogenetic group (*12*). Since 1918, seasonal IAV outbreaks have been caused by HA group 1 subtypes H1 and H2, and HA group 2 subtype H3. The 1918-1920 “Spanish flu” pandemic was an integral event in the history of human exposure to IAV H1N1, with an estimated global death toll of over 15 million (*13*). Phylogenetic and seroarcheological testing have provided a plausible hypothesis that H1 was first transmitted to humans from birds before 1907, likely as H1N8, and N1 was later transmitted from birds around 1915 once H1 was already established in humans (*6, 14, 15*). RNA recovered from preserved tissue samples of 1918 influenza victims confirmed that the structure and sequence of the 1918 H1N1 virus resemble those of avian strains (*15, 16*).

The antigenic evolution of H1N1 in the first half of the twentieth century can be divided into several distinct phases, beginning with the 1918 pandemic, with influential changes occurring in the early 1930s and mid-1940s (*1, 6, 17–19*). Classical swine H1N1, the IAV lineage first isolated from hogs in 1930, shares antigenic characteristics with the 1918 pandemic human virus and human H1N1 viruses isolated in 1933 (*14,18,20–23*). H1N1 strains isolated between 1934 and 1943 diverge from the 1918 virus, but appear related to each other, forming the “A” lineage (*1, 18*). During the 1946-1947 influenza season, H1N1 underwent further antigenic change, resulting in the “A-Prime” variant that circulated until 1957 (*1, 24, 25*). These transitions define at least three distinct antigenic phases of H1N1 circulation – H1N1_*⍺*_ (1918 –early 1930s), H1N1_*β*_ (mid-1930s–early-1940s), and H1N1_*γ*_ (mid 1940s–1957).

In 1957, major antigenic shift in circulating IAVs led to the introduction of H2N2 in human populations, triggering a pandemic as H2N2 overtook the vast majority of influenza infections. H2N2 circulated for ten years until extinction during the 1968 influenza pandemic, when another antigenic shift introduced H3N2, marking the transition from a group 1 IAV dominated era to a group 2 IAV dominated era (*2,26,27*). Since their 1968 emergence, H3N2 viruses have evolved extensively and rapidly, demonstrating the highest rate of antigenic drift among all human IAVs and causing large seasonal outbreaks with substantial mortality (*28–30*). H1N1 reentered human populations in 1977 and has since remained in seasonal co-circulation with H3N2 (*31*). In 2009, a novel H1N1 virus, H1N1pdm09, was detected in North America and quickly spread globally, replacing seasonal H1N1 strains that had circulated since the late 1970s (*32*). This pandemic influenza virus most closely resembles the earlier pandemic H1N1 variant, pH1N1_*⍺*_, which circulated between 1918 and the early 1930s (*33*).

Since H1N1 was the only influenza strain in circulation between 1918 and 1957, birth cohorts spanning this period may have all been imprinted with related H1N1 strains. These cohorts’ immune responses to later-life influenza exposures may vary depending on the antigenic characteristics of the H1N1 strain they were exposed to in childhood. Although the age pattern of mortality during the 2009 H1N1pdm09 pandemic is well documented (*2, 7, 34–37*), mortality differences across cohorts first exposed to each of the three distinct H1N1 evolutionary phases have not yet been fully explored. Many open questions also remain about how imprinting shapes a cohort’s influenza mortality risk across their lifecourse, and how population-level trends in influenza mortality are shaped by the complex interactions between childhood immune imprinting and the virulence of the circulating seasonal strain.

We analyzed U.S seasonal influenza mortality by age and single-year birth cohort to disentangle the effects of age, the circulating strain, and childhood immune imprinting. To accomplish this, we first decompose mortality trends into age-specific and period-specific components, using a novel application of a common mortality forecasting method. This framework is useful for studying immunity and infection dynamics for diseases such as influenza, which exhibit strong age-gradients and period effects in severity and mortality due to many factors including the circulating strain and the population immune landscape. Using a statistical model, we then quantified the relative risk of seasonal influenza mortality experienced by cohorts imprinted with different strains in childhood. We find large differences in mortality risk over a cohort’s lifetime based on the imprinted strain. Our results highlight the long-term population-level consequences of childhood immune imprinting, and potential implications for the future dynamics of influenza mortality.

## Materials and Methods

### Constructing a panel of influenza surveillance data and mortality data

We constructed single-year age-specific influenza mortality rates for all birth cohorts born between 1860 and 2020 using individual-level mortality data from the National Center for Health Statistics (NCHS) National Vital Statistics System multiple cause of death data series (*38*) for the years 1968-2021. NCHS mortality data are derived from every death certificate filed in the US each year (data for 1972 only include a 50% sample, which has been weighted for our analysis). The data include an underlying cause of death (UCOD) and up to twenty ancillary causes of death, indicated by three versions of ICD codes: ICD-8 (1968-1978), ICD-9 (1979-1998), and ICD-10 (1999-2021). We defined an influenza-attributable death as a death in which an ICD code for influenza (ICD-8: 470–474; ICD-9: 487–488; ICD-10: J09–J11) appears anywhere in the death record. All influenza-related deaths were included in our analysis. Individual records of death were aggregated by birth year and influenza season (defined below) based on month of death. We estimated birth years by subtracting age of death from calendar year of death, and assigned deaths to influenza seasons using the exact month of death.

We defined an influenza season as starting in September of one year and ending in August of the following year. Although the 2009 influenza pandemic began earlier in the spring (H1N1pdm09 was first detected in the US in late April 2009 (*32*)), we chose to use the same definition of a season for all years included in our study in order to make mortality rates comparable across seasons. We calculated mortality rates for single-year age groups for each influenza season by dividing the number of deaths by single-year age and single-year person-years of exposure. The number of person-years of exposure over the course of an influenza season was estimated from the Human Mortality Database (HMD) (*39*) annual exposure data, which provides annual estimates as of January 1st. We interpolated adjacent yearly values to derive monthly estimates; from this, we extracted exposure for September of each year to approximate the population at risk at the start of each influenza season.

We classified each influenza season from 1968-1969 to 2020-2021 as H1N1-, H1N1pdm09-, H3N2-, or B-dominant if a single strain accounted for 70 percent or more of positive virus specimen test results; otherwise, we classified seasons as ambiguous. The dominant circulating strain for each season are provided in Supplementary table S2. Specimen results for the years 1968-1998 were compiled from the CDC’s archive of influenza surveillance reports (*40*). For 1997-2021, specimen test data were obtained from the WHO’s FluNet (*41*) and restricted to reports from U.S.-based testing facilities. These data are limited by their dependence on accurate reporting by the collaborating laboratories. Additionally, milder or undiagnosed influenza cases may not undergo testing, which could skew results toward viruses that cause more severe infections. Among the influenza seasons classified using FluNet data, the 2020-2021 season was the only one in which influenza B specimens (of either lineage) outnumbered all combined IAV subtypes. As a result, we classified this period as the only B-dominant season of the twenty-first century. For all other FluNet-derived seasons, we determined the dominant strain by using only specimen results for IAV subtypes.

### Assigning imprinted strains by birth year

Following the methods of Gostic *et al.* (2016) (*3*) (*42*), we assigned probabilities of being imprinted by H1N1, H3N2, and H2N2 to each birth cohort born after 1917. Cohorts born prior to 1918 were labeled as “ambiguous”. For the 1918 and beyond cohorts, an imprinting probability of 70 percent or higher was required to assign an imprinted subtype. Cohorts that did not have at least 70 percent probability of having been imprinted by a particular subtype were labeled as “ambiguous.” These ambiguous cohorts make up 27 percent (n = 26 out of 98) of the 1918 and beyond cohorts. Young children have a high probability of being immunologically naïve to IAV (*3*). Because of this, birth cohorts born during or shortly before the study period were typically classified as naïve during influenza seasons in their early childhood. These cohorts were assigned an imprinted virus or labeled as ambiguous (following our 70 percent threshold) once the probability of naïvete was lower than the probability of imprinting to any virus. Ambiguous cohorts could be reclassified as imprinted in later seasons if a virus reached 70 percent probability of imprinting. Cohorts born between 1918 and 1954, before the 1957 H2N2 pandemic, were all likely imprinted with H1N1. H1N1 cohorts were further subdivided to account for the antigenic evolution of the virus in the early twentieth century: novel pandemic-H1N1_*⍺*_, (1918-1920), seasonal H1N1_*⍺*_ (1921-1932), H1N1_*β*_ (1933-1946), and H1N1_*γ*_ (1947-1953). We modeled imprinting for the H1N1 subtypes purely based on being born between the year that major antigenic change occurred and the end of a variants time in circulation. The most probable imprinted strains by birth year are provided in Supplementary table S3.

### Mortality fitting with the Lee-Carter model

To characterize mortality differentials between cohorts imprinted by different influenza strains, while accounting for age- and period-effects, we modeled influenza mortality rates following the formulation of Lee and Carter (*43*). The Lee-Carter model is one of the most widely-used approaches for the stochastic modeling and forecasting of age-specific mortality rates. Using a logit-link extension of the Lee-Carter model, we modeled age-specific period mortality rates as a function of three components:

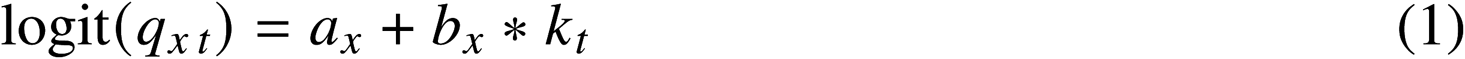

where *q*_*x*,*t*_ is the mortality probability for single-year age *x* at single-period time *t*, *a*_*x*_ captures the average log influenza mortality rate for age *x* across all periods, *k*_*t*_ is a time-varying index of the level of mortality at time *t*, and *b*_*x*_ measures the sensitivity of mortality at age *x* to changes in *k*_*t*_. In this framework, the age effect, *a*_*x*_, is the time-invariant baseline age-shape of mortality (static age function). Temporal changes in mortality are entirely captured by *k*_*t*_, which is shared across ages. *b*_*x*_ acts as a weight to control how each age is affected by temporal mortality fluctuations. This specification assumes that the age-dependent mortality pattern is independent of birth year. We estimated the parameters of the Lee-Carter model using maximum likelihood estimation, and assuming that the probability of dying follows a binomial distribution (*44*). To construct confidence intervals for the model parameters and fitted mortality rates, we implemented a semi-parametric bootstrap procedure (*45, 46*). Death counts were resampled with replacement to refit the model 1600 times and obtain 1600 estimates each of *a*_*x*_, *b*_*x*_, and *k*_*t*_. We computed upper and lower bounds at the 95 percent confidence level based on the distribution of the parameter estimates, and the distribution of *q*_*x*,*t*_ computed for each replication.

In our application, the Lee-Carter model tests whether the age-shape of influenza mortality is constant over time, with only the overall level of mortality fluctuating season to season. In other words, the model tests the null hypothesis that differences in mortality are primarily age-based. Because all cohorts pass through the same ages at different times, birth-year-specific immune histories do not alter the age-specific parameters, *a*_*x*_ and *b*_*x*_. Therefore, discrepancies between in-sample observed mortality and Lee-Carter fitted mortality may indicate a birth-year- and period-specific effect not captured by the age terms. As a sensitivity test, we fit our data to the Cairns-Blake-Dowd (CBD) model (*47*), another widely-used stochastic mortality model that forgoes a cohort effect term. The Lee-Carter model performed exceptionally better than the CBD model at fitting seasonal influenza mortality (Supplementary figure S4; supplementary table S4). We also fit four additional stochastic mortality models that include a cohort term: The Renshaw-Haberman model (*48*), Age-Period-Cohort (APC) model (*49*), Extended CBD model (*49*), and the Plat model (*50*) (see Supplementary table S4 for formulation details). We additionally refit the Lee-Carter model separately with data prior to 2009, and after 2009, to test sensitivity of the over-saturation of H3N2-dominated seasons in the study period.

We calculated the deviance residuals of the fitted mortality models to quantify how observed mortality deviated from the expected pattern. Because the original Lee-Carter framework captures age- and period-specific trends, and omits a cohort effect, we assume that residual deviance in the fitted Lee-Carter model signals a mortality effect that is dependent on period and birth-year. In other words, when modeled age-specific mortality rates fail to align with observed data for certain cohort-season pairings, the discrepancy may reflect cohort-specific risks driven by shared immune histories. In line with immunological intuition, we hypothesized that a birth cohort’s mortality rates would be lower than our model predicted in seasons dominated by the virus subtype a cohort was most exposed to in early childhood. For instance, cohorts first exposed to H1N1 would have lower mortality than expected during H1N1-dominant years. On the other hand, if imprinting protection works at a narrower and more specific scale, we would expect cohorts imprinted with H1N1 variants most related to the original 1918 pandemic virus (H1N1_*⍺*_) to be more protected from mortality due to H1N1pdm09 compared to those imprinted with later, less similar H1N1 variants. If imprinting provides broad HA group-level protection against seasonal influenza mortality, then we would expect cohorts first exposed to group 1 HA subtypes H1 or H2 to experience lower mortality risk during H1N1-dominant years, and cohorts first exposed to group 2 H3 to experience lower mortality risk during H3N2-dominant seasons.

### Statistical model for the interaction between imprinted strain and circulating strain

Finally, we directly test the interaction effect between a cohort’s imprinted strain and the dominant circulating strain on seasonal influenza mortality. To do so, we fit a negative binomial regression model to estimate the associations between age-specific seasonal influenza mortality and (1) cohorts’ most likely imprinted strains, (2) the dominant circulating strain, and (3) the interaction between imprinted strain and the circulating seasonal strain. We also included interactions between the circulating strain and cumulative exposure to H1N1, H2N2, and H3N2, a natural spline of age, and seasonal fixed effects. We used the negative binomial framework to account for over-dispersion in mortality counts. We estimated the following model:

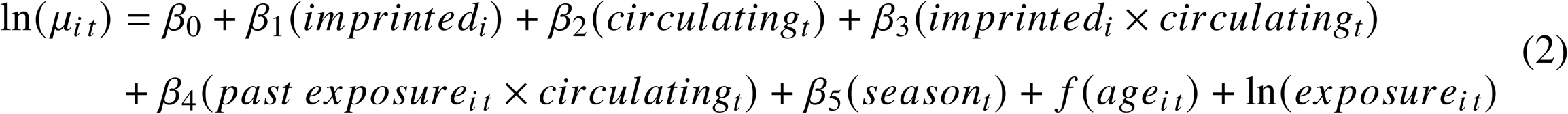

where *μ*_*i* *t*_ is the count of deaths for birth-cohort *i* in time period *t*, *imprinted*_*i*_is a categorical variable indicating the strain a birth-cohort could be imprinted with: pH1N1_*⍺*_, H1N1_*⍺*_, H1N1_*β*_, H1N1_*γ*_, H2N2, H3N2, mixed strains, unknown strain (born prior to 1918), or naive; *circulating*_*t*_indicates whether influenza B, H3N2, H1N1, H1N1pdm09, or an ambiguous strain is the dominant strain circulating in a season; (*past exposure*_*i* *t*_ is the cumulative sum of seasons in the study period that each cohort was exposed to either H1N1, H2N2, or H3N2, normalized by total influenza seasons survived; *f* (*age*_*i* *t*_) is a natural spline with 8 degrees of freedom; *season*_*t*_ is a categorical variable indicating the time period; and the offset term is *ln*(*exposure*_*i* *t*_), the natural log of person-years of exposure to the risk of dying from influenza. In the results presented here, the *imprinted*_*i*_ reference level is an H3N2-imprinted cohort, and the *circulating*_*t*_ reference level is an H3N2-dominated season.

The model includes an interaction between a cohort’s imprinted strain and a season’s dominant strain to capture the joint effect on mortality. This allows us to determine to what degree imprinting to a specific variant affects strain-specific mortality outcomes for each cohort. We modeled age as a natural spline to avoid the assumption of a linear functional form between mortality and age. While imprinted cohort and age are highly correlated within any given influenza season, our analysis spans all seasons from 1968-1969 to 2019-2020. This temporal coverage allows us to observe the same imprinted cohorts at different ages across all seasons, meaning each age contains individuals with different immune histories. We include cumulative exposure to H1N1, H2N2, and H3N2 in order to distinguish the effect of initial strain imprinting from any additional protection caused by repeat exposure. For cohorts born within the study period, cumulative exposure to all strains begins at 0 and increases sequentially for each season that H1N1 or H3N2 was the dominant strain (H2N2 disappeared in human populations in 1968). For cohorts born before the study period, we calculated exposure prior to 1968 from the years lived during circulation of H1N1 (1918-1957) and H2N2 (1957-1968). We report the rate ratios (RR) estimated from the negative binomial regression, and the associated 95% confidence intervals.

## Results

### Decomposing influenza mortality trends into age- and period-specific components

We constructed a panel of age-specific influenza mortality rates for U.S. birth cohorts born between 1860 and 2020, computing each cohort’s single-season and single-age rates for every influenza season between 1968-1969 and 2020-2021. We classified each influenza season as H1N1-, H1N1pdm09-, H3N2-, or B-dominant if that strain accounted for at least 70 percent of positive virus specimens in that season; otherwise, we classified the season as ambiguous. Following the methods of Gostic *et al.* (2016) (*3*), we assigned probabilities of being imprinted with H1N1, H2N2, and H3N2 to each birth cohort born after 1917. H1N1 cohorts were further subdivided to account for the antigenic evolution of the virus in the early twentieth century: novel pandemic-H1N1_*⍺*_, (1918-1920), seasonal H1N1_*⍺*_ (1921-1932), H1N1_*β*_ (1933-1946), and H1N1_*γ*_ (1947-1953).

The history of influenza virus circulation has led to notable differences in cohorts’ influenza mortality profiles over their lifescourse. Figure 1 summarizes the influenza mortality experience of cohorts imprinted with different IAV strains over time (Figure 1A) and by age (Figure 1B). For all cohorts, we see a dip in mean influenza mortality corresponding to declining levels of overall seasonal influenza mortality starting in the mid-1970s (Fig. 2D). As expected, younger cohorts imprinted with H2N2 and H3N2, have lower mean mortality throughout the study period, compared to older H1N1 imprinted cohorts, corresponding with the well-established age-gradient in influenza mortality. Strikingly, we see no significant mortality differences between the youngest H3N2 cohorts and the older, H2N2 cohorts until the late 2000s. This is most evident when we summarize mortality rates by age (Figure 1B). For H3N2-imprinted cohorts born between 1968 and 1976, on average, 6.04 per 100,000 of each birth cohort died from an influenza-related death between the ages of 12 and 45 (95% CI: 5.49–6.59). In contrast, a much smaller share, 1.84 per 100,000 (95% CI: 1.65–2.03), of each H2N2-imprinted birth cohort died from an influenza-related cause between the same ages, on average. We find that cohorts first exposed to different strains exhibit distinct influenza mortality age profiles over their lifetime. The younger H3N2-imprinted cohorts have experienced substantially higher age-specific mortality rates when compared to the experience of older cohorts at the same ages.

**Figure 1:**
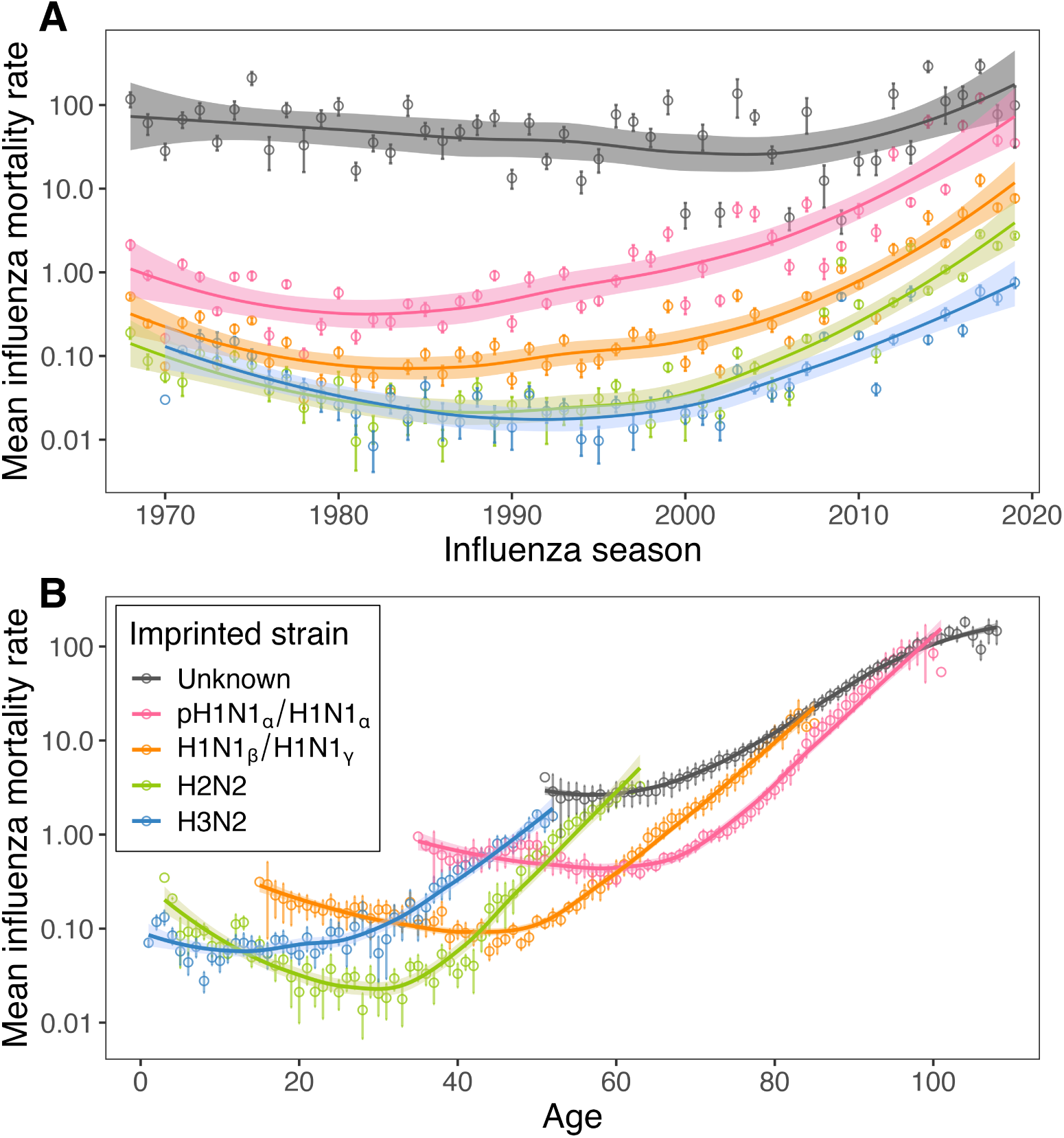
Observed period and cohort influenza mortality rates. (A) Observed age-specific influenza mortality rates averaged across birth cohorts imprinted with the same strains in all influenza seasons between 1968-1969 and 2019-2020. (B) Observed period influenza mortality rates for birth cohorts imprinted with the same strains, averaged across all ages each imprinted cohort spans each season. (A-B) Smooth line and confidence intervals are derived from LOESS regression. Error bars around the observed points represent the mean mortality rate ± the standard error of the mean. The absence of error bars around a point indicates that only one observation was available for that imprinted group at that age or season. The mortality rates are calculated per 100,000 population and plotted on a log scale.

**Figure 2:**
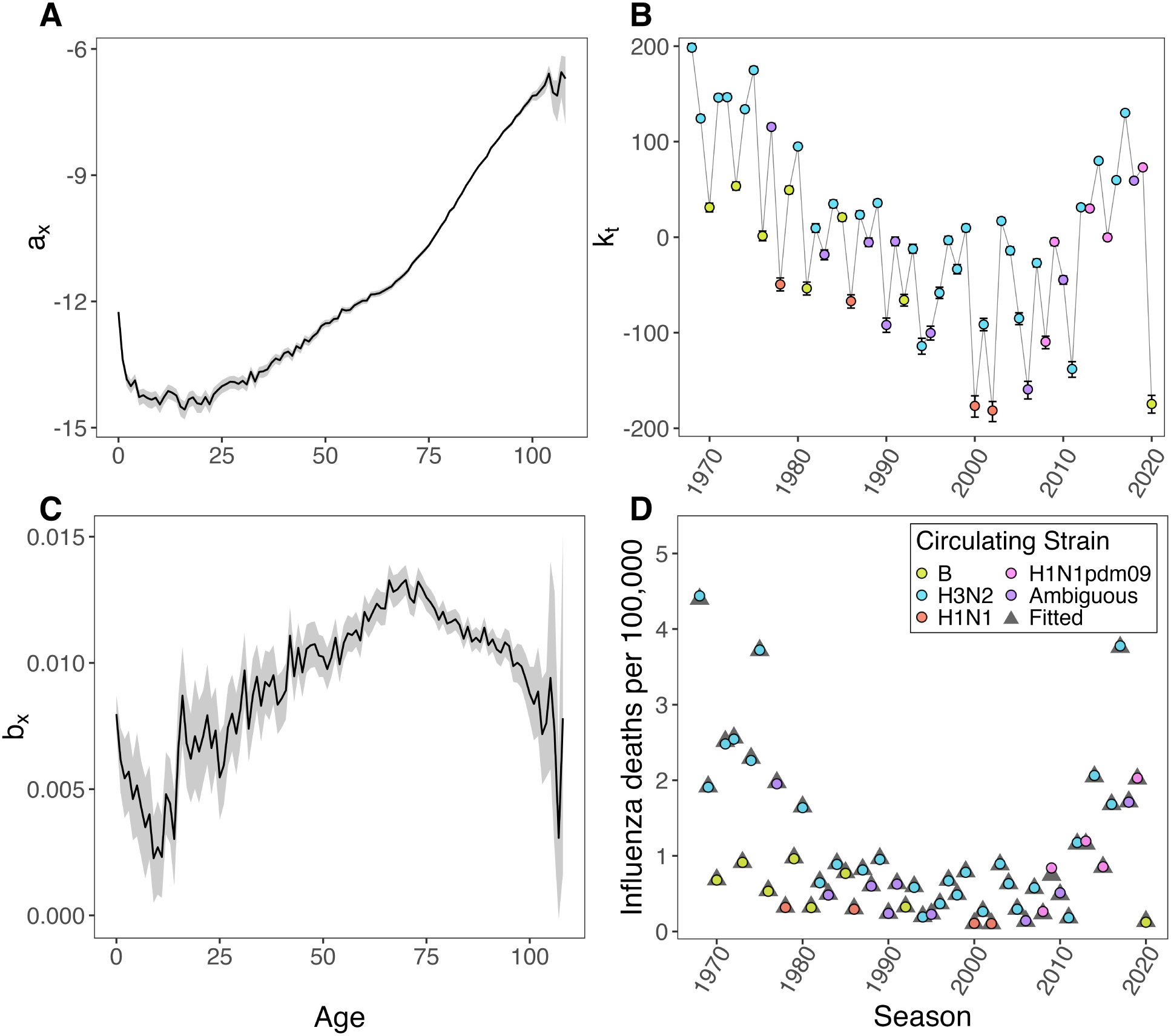
Lee-Carter fitted model parameters and observed period influenza mortality rates. (A-C) Lee-Carter estimates and bootstrapped 95% confidence intervals for (A) *a*_*x*_, which captures the time-invariant average log influenza mortality rate for each age across all periods; (B) *k*_*t*_, the time-varying index of the level of mortality across time; (C) *b*_*x*_, which captures how each age is affected by temporal mortality fluctuations. (D) Observed and fitted total influenza mortality rate per 100,000 in each season.

To understand these cohort-period differences, we decomposed influenza mortality over the past 53 influenza seasons into age- and period-specific components using a framework commonly used to forecast mortality – the Lee-Carter model. The Lee-Carter model decomposes observed age-specific mortality into three components – time-averaged age-specific mortality profile, *a*_*x*_; a time-series vector, *k*_*t*_, capturing variations in overall mortality over time; and a vector *b*_*x*_ that describes the responsiveness of mortality in each age group to overall temporal changes in mortality.

Influenza mortality is strongly associated with age, as evident from the age parameter estimates from the fitted Lee-Carter model, *a*_*x*_, which captures the log age-shape of influenza mortality (Fig. 2A) averaged over the observed time period. As expected, we find the typical “U-shaped” age-profile, indicating higher mortality rates for very young children in comparison to children at older ages. The heightened mortality rate at young ages decreases into adolescence, then increases continuously with age. The vector, *b*_*x*_(Fig. 2C), captures how sensitive each age is to overall temporal fluctuations in influenza mortality. Higher values of *b*_*x*_ indicate ages at which mortality changes the most rapidly in response to overall changes in mortality, reaching a peak at age 70.

Figure 2B shows overall temporal trends in influenza mortality as estimated by the temporal index parameter *k*_*t*_. Fluctuations in *k*_*t*_ are largely consistent with strain-specific variation in mortality and morbidity observed season to season. As has been described in previous work, and confirmed by our regression results discussed below, seasons dominated by H3N2 have higher mortality at all ages, while H1N1- and influenza B-dominated seasons generally have lower mortality (*51, 52*). Our estimates for *k*_*t*_ shows a general decline in influenza mortality between 1968 and 2009, with an increase following the 2009 pandemic, consistent with higher mortality in H1N1pdm09-dominated seasons compared to pre-2009 H1N1 seasons. A sharp drop in overall mortality is evident for the 2020-2021 season, a period that was characterized by COVID-19 related use of non-pharmaceutical interventions (*53*). The Lee-Carter model estimates, aggregated by season, closely correspond to total observed seasonal influenza mortality (Fig. 2D). Notably, the upward trend in influenza mortality since the 2010s, shown in figure 2D, may be indicative of increasing severity in recent influenza seasons. This trend may also be due to changes in diagnostic practices and death certificate coding, as well as population aging increasing the proportion of the population vulnerable to influenza mortality, as nearly half of all influenza deaths occurring in the U.S. in this period were aged 85 years and older (*54*).

### Deviations from Lee-Carter model fit reveal cohort-period interaction effects on influenza mortality

To identify if certain cohorts, age groups, or periods have higher or lower mortality than expected based on historical trends, we calculated the residuals between observed and expected mortality, based on the model fit, for each season and age group. The Lee-Carter deviance residuals plotted on an age-period surface (Fig. 3) reveal a striking difference in the age-pattern of residuals before and after 2009. We find that since 2009, in H1N1pdm09-dominated seasons, deviance residuals between expected and observed mortality show a distinct age pattern: older cohorts exhibit lower mortality compared to modeled rates (negative residual deviance), while younger cohorts cross over into excess mortality (positive residual deviance). The age-pattern in mortality residuals is most pronounced for the 2009 pandemic season, but the 2013-2014, 2015-2016, and 2019-2020 H1N1pdm09-dominant seasons also follow a similar age pattern.

**Figure 3:**
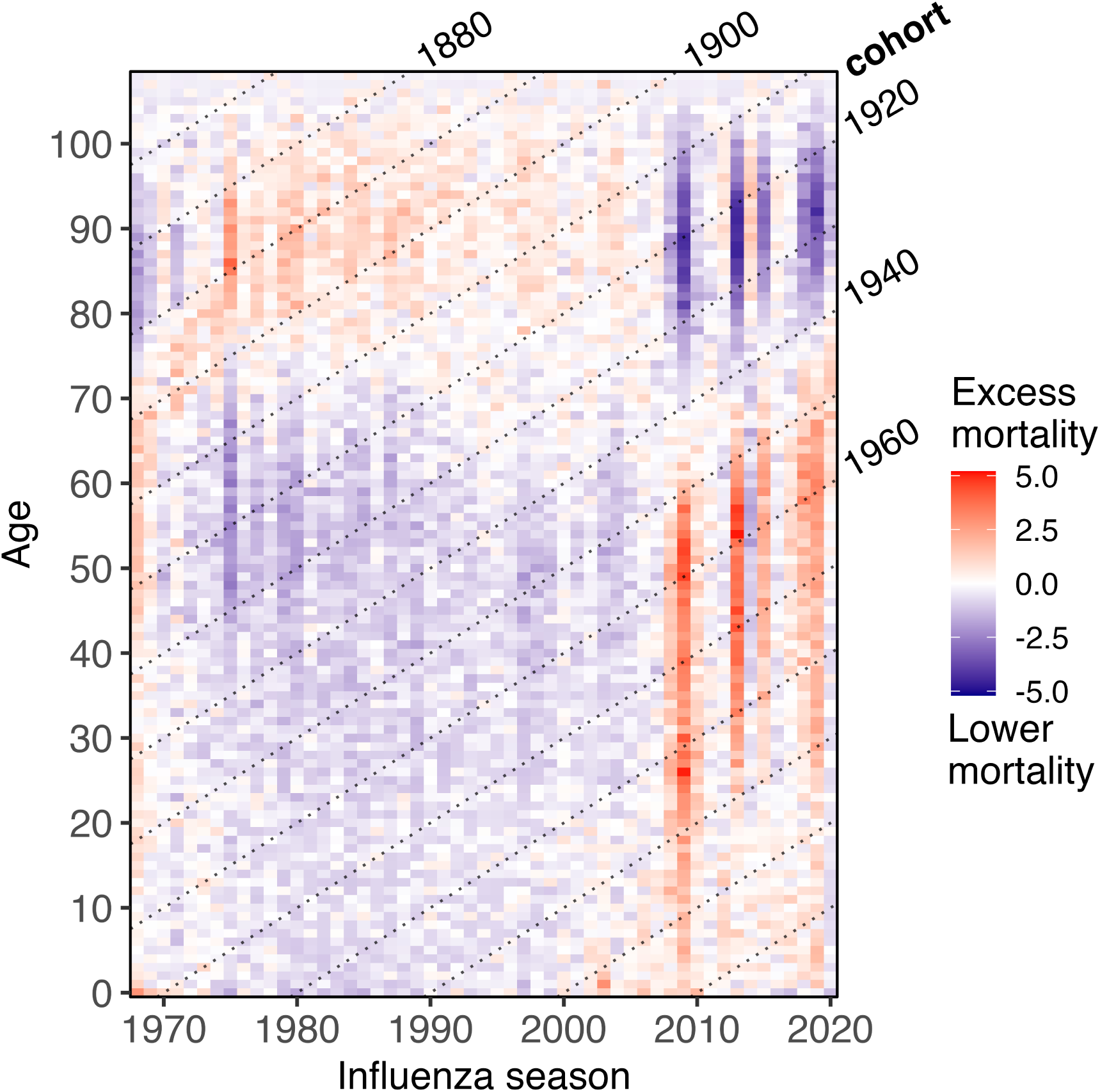
Deviance residuals of the Lee-Carter model by age and influenza season. Each cell represents a single-year age and single influenza season. The x-axis labels corresponds to the influenza season. Residuals are adjusted to account for overdispersion. Positive residuals (in red) indicate higher observed age-specific mortality rates in comparison to the Lee-Carter fitted mortality rates; negative residuals (in blue) indicate lower observed mortality than estimated.

Notably, we find no similar residual patterns when we replicate our analysis to fit all-cause mortality (Supplementary fig. S1) and mortality from pancreatic cancer (Supplementary fig. S2). This suggests that residuals in influenza mortality are not solely caused by an overall period effect. The residual patterns from the Lee-Carter model fit also do not appear to be a pure birth cohort effect, which would be the case if certain birth cohorts had a mortality advantage or disadvantage throughout their lives. These would appear as diagonal bands in the residual structure of influenza mortality. As a sensitivity test, we fit four additional stochastic mortality models formulated to capture cohort effects (Supplementary table S4). Cohort models outperform the simpler Lee-Carter model. However, the residual structure remained in most models (Supplementary figs. S3-S4), suggesting that a birth cohort effect alone cannot explain the observed pattern. Since the vast majority of years prior to 2009 were dominated by H3N2, we also refit the Lee-Carter model separately to mortality data prior to 2009 and to data after 2009; these model fits yield a visually similar residual structure (Supplementary fig. S5), suggesting that these results are not an artifact of the abundance of H3N2 dominated seasons relative to seasons dominated by other strains.

Figure 4 illustrates the model’s fit to in-sample data, by age and birth year, for select influenza seasons dominated by different influenza strains prior to 2009 (Supplementary Figs. S6-S12 show the model fit for all seasons in the study). Discrepancies in modeled mortality rates prior to 2009 appear as over-estimated mortality in younger ages, while older adults’ fitted mortality rates align closely. Observed mortality during seasons dominated by H3N2 or Influenza B, or H1N1 prior to 2009, or those without a dominant strain, do not show any significant deviations from fitted rates.

**Figure 4:**
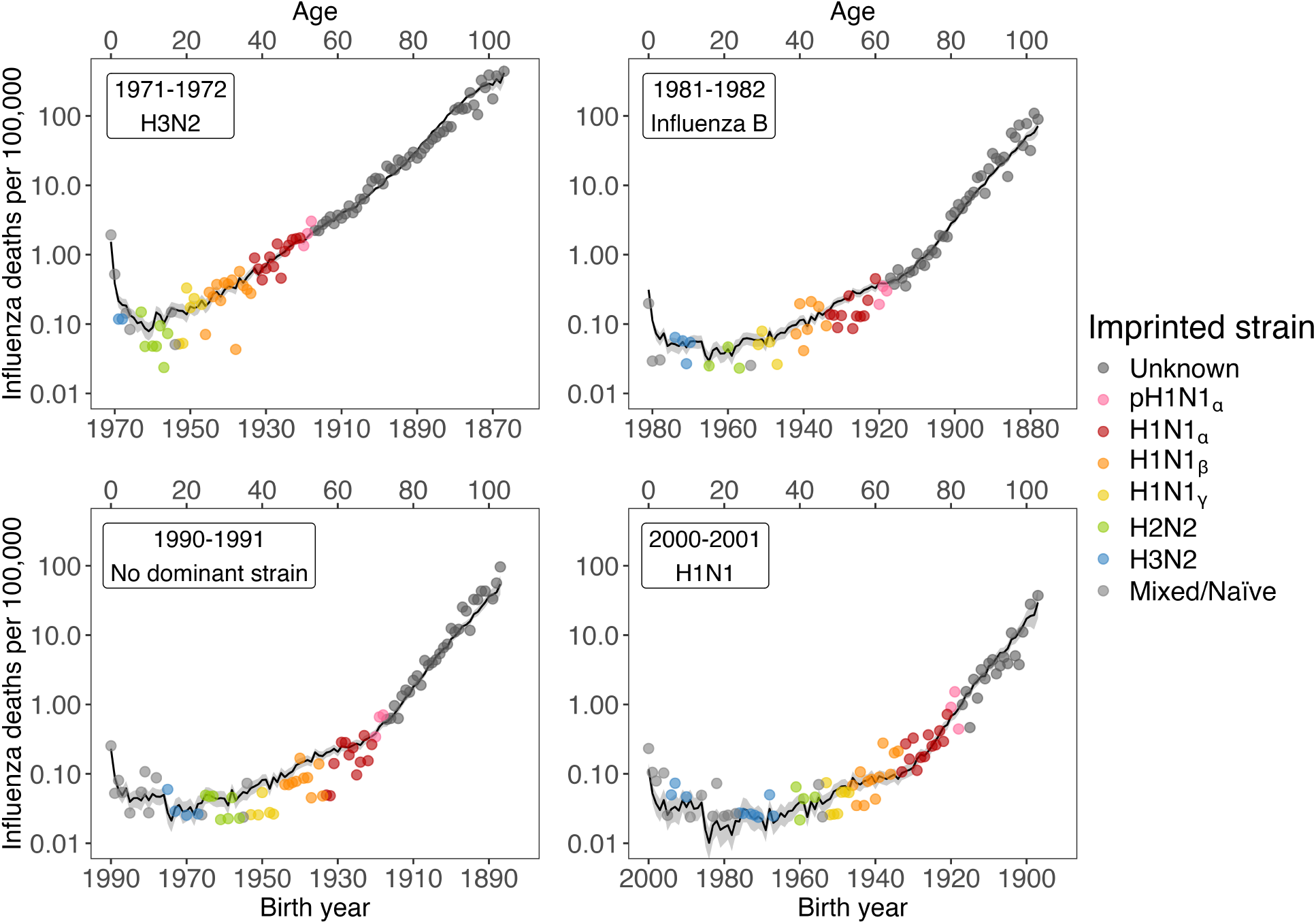
Lee-Carter fitted influenza mortality rates per 100,000 compared to observed rates, for select influenza seasons before 2009. Supplementary Figs. S6-S12 show the model fit for all seasons in the study. Solid black line represents the fitted rates with 95 percent confidence intervals, and each point represents the in-sample observed single-year age-specific mortality rate for a birth cohort. Rates are plotted on a log scale.

Figure 5 shows in-sample deviations in influenza mortality for select influenza seasons since 2009 (Supplementary Figs. S6-S12 show the model fit for all seasons in the study). For all H1N1pdm09-dominant seasons, the residual crossover happens around the 1940 to 1944 birth cohorts. These cohorts encountered the earliest H1N1 strains (H1N1_*⍺*_ and older H1N1_*β*_ cohorts) in childhood. This aligns with the hypothesis that antigenic similarities between the 1918 and 2009 H1N1 viruses offer protection against H1N1pdm09 for individuals that have been exposed to the 1918 virus. Although we observe a consistent crossover at cohorts born in the early 1940s, this transition may not suggest a single change point based on imprinted strains. The gradual transition between positive and negative deviance could reflect cumulative exposures to early H1N1 strains, with older cohorts having sequentially more opportunities for reinforced immunity. This may also explain why cohorts born prior to 1918, whose first exposures were to pre-pandemic strains, do not follow the same deviation. Finally, excess mortality among H2N2 cohorts than would be expected for their age-cohort suggests a lack of HA group-level imprinting protection. Overall, the difference in deviance residuals, between the pre-2009 and post-2009 era suggest that these patterns may be caused by an unmodeled cohort-period interaction effect, such as an interaction between cohorts’ imprinted strains and seasonal circulating strains. Next, we test this hypothesis directly using a statistical model described below.

**Figure 5:**
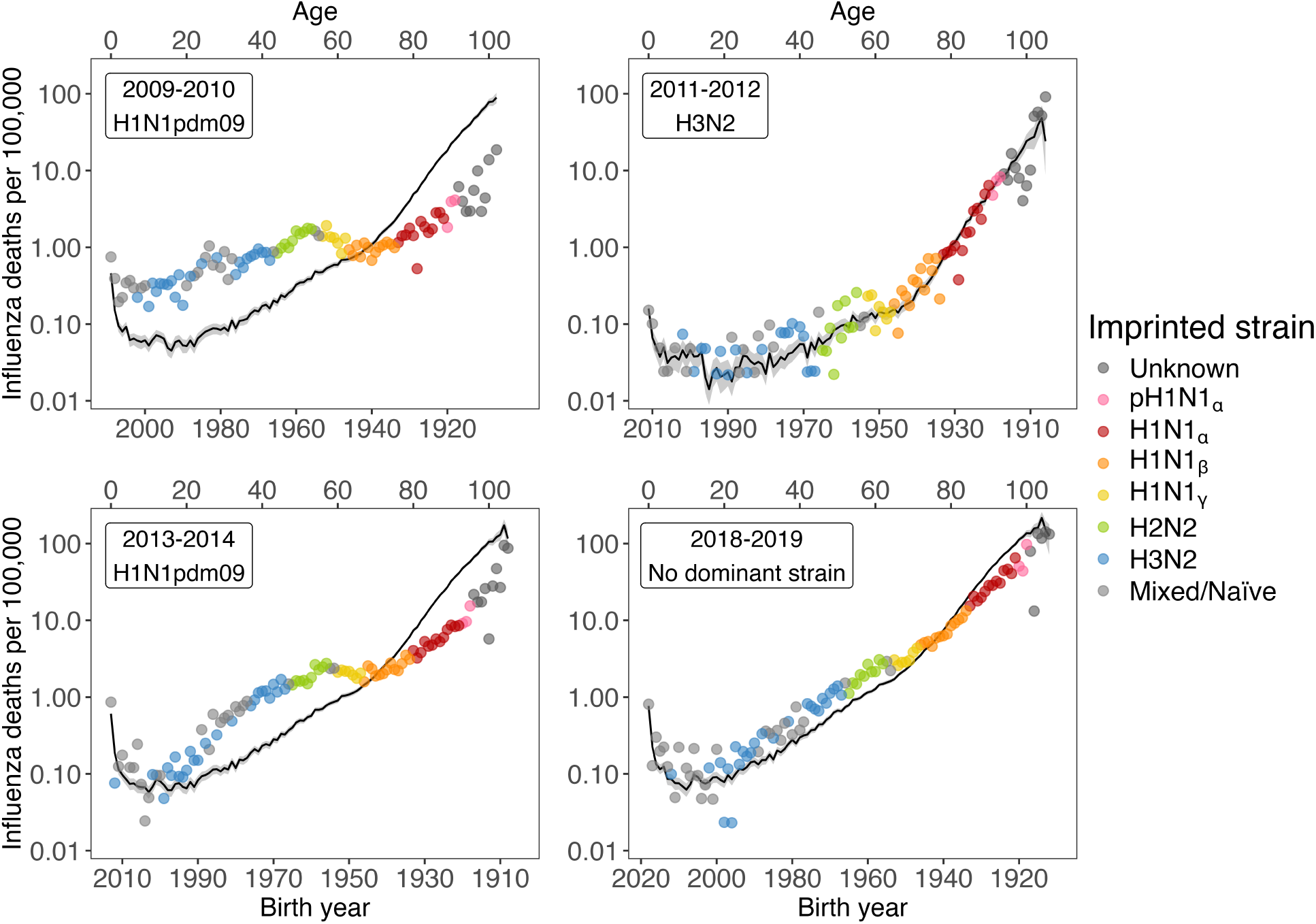
Lee-Carter fitted influenza mortality rates per 100,000 compared to observed rates, for select influenza seasons since 2009. Supplementary Figs. S6-S12 show the model fit for all seasons in the study. Solid black line represents the fitted rates with 95 percent confidence intervals, and each point represents the in-sample observed single-year age-specific mortality rate for a birth cohort. Rates are plotted on a log scale.

### Relative mortality differences by circulating strain and imprinted strain

To directly test the interaction effect of a cohort’s imprinted strain and the dominant strain circulating in a season, we use a negative binomial model to estimate relative mortality differentials by circulating strain and cohorts’ imprinted strains, adjusting for age, period, and cumulative exposure to the circulating strain. Evidence of immune protection consistent with imprinting patterns are reflected in our results from the negative binomial model (Supplementary table S1). Figure 6 is a marginal effects plot showing regression adjusted influenza mortality rates per 100,000 (in log scale) for each imprinted cohort across seasons with different dominant strains. To visualize the interaction effect between the circulating strain in a season and the imprinted strain of cohorts alive during that season, we plot the adjusted influenza mortality rates per 100,000, varying the circulating and imprinted strain, but holding season, age and cumulative exposure constant. In line with previous research, we find that, holding all covariates constant, H1N1-dominated seasons prior to 2009 were associated with significantly lower mortality rates overall compared to H3N2-dominated seasons (RR: 0.031, 95% CI: 0.022, 0.052). While H1N1 mortality rose after H1N1pdm09 replaced previous H1N1 strains, this increase is not statistically significant relative to H3N2 seasons. Notably, we find limited evidence of imprinting protection among H3N2-imprinted cohorts during H3N2-dominated seasons. In fact, only cohorts born prior to 1918 experienced higher mortality than H3N2-imprinted cohorts during H3N2 seasons, after adjusting for age (RR: 1.398, 95% CI: 1.175, 1.663). Further, cohorts imprinted with H1N1_*β*_, H1N1_*γ*_, and H2N2 all exhibited lower mortality than H3N2 cohorts during H3N2 seasons, after adjusting for age (RR: 0.807, 0.807, 0.780, respectively). No significant mortality differences were found between H3N2-imprinted cohorts and all other imprinted groups during H3N2 seasons.

**Figure 6:**
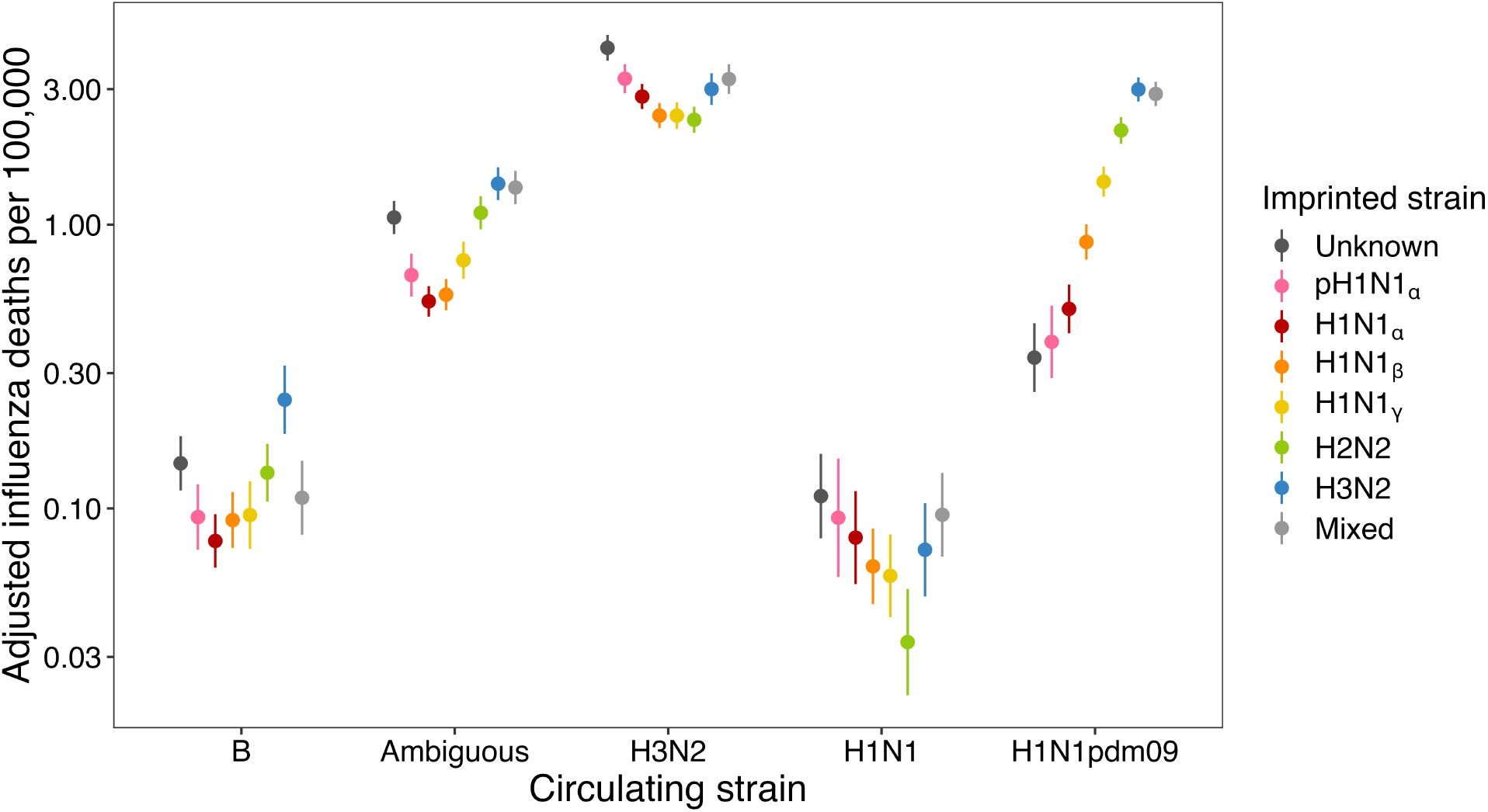
Adjusted influenza mortality rates from a multivariate negative binomial regression model fit to observed influenza mortality data. Mortality rates are shown per 100,000 and plotted on a log scale. The model includes age splines, categorical variables for each season, and a control for cumulative exposure to the circulating strain. The predicted mortality rates shown are marginal effects, i.e. they are based on the estimated interaction effect between imprinted strain and circulating strain, while holding season at the reference level (1968-1969), and age and cumulative exposure at their mean values (age: 55.1; cumulative exposure: 19.5% of survived seasons dominated by the circulating strain).

Before the emergence of H1N1pdm09, H1N1-imprinted cohorts exhibited no significant mortality differences relative to H3N2-imprinted cohorts during H1N1-dominated seasons. However, we find notable differences in mortality rates starting in 2009. All H1N1-imprinted cohorts consistently experienced significantly lower mortality during H1N1pdm09-dominated seasons compared to H3N2-imprinted cohorts. In H1N1pdm09 seasons, the lowest estimated mortality was seen for cohorts imprinted with pH1N1_*⍺*_ (RR: 0.119, 95% CI: 0.050, 0.279), after adjusting for age, followed by H1N1_*⍺*_ (RR: 0.179, 95% CI: 0.092, 0.347), H1N1_*β*_ (RR: 0.359, 95% CI: 0.241, 0.535), and H1N1_*γ*_ (RR: 0.586, 95% CI: 0.488, 0.705). These results suggest enhanced protection against H1N1pdm09 for older cohorts first exposed to 1918 pandemic H1N1_*⍺*_ or seasonal H1N1_*⍺*_ viruses, and progressively weaker protection for cohorts imprinted with later variants of H1N1. This enhanced protection may have come at a cost, as pandemic and seasonal H1N1_*⍺*_-imprinted cohorts experienced relatively higher mortality in H3N2-dominated seasons compared to younger cohorts, even after adjusting for age.

These results shed light on the observed differences in age-specific mortality rates experienced by cohorts imprinted with different strains (Fig. 1). The younger H3N2-imprinted cohorts have experienced substantially higher age-specific mortality rates when compared to the experience of older cohorts at the same ages (Fig. 1B). This has been due to the emergence of H1N1pdm09, and the relative age-adjusted mortality risk for H3N2-imprinted cohorts compared to older cohorts. Using our fitted negative binomial model, we can estimate the mortality burden for H3N2- and H2N2-imprinted cohorts in future H1N1pdm09-dominated seasons by estimating the influenza mortality rate for these cohorts at an older age. Holding age at 65, season at 2019-2020 (the most recent H1N1pdm09 season in our dataset), and cumulative exposure at the mean, we project 6.47 influenza deaths per 100,000 H3N2-imprinted individuals in each future H1N1pdm09 season (95% CI: 5.84–7.167), and 4.64 deaths per 100,000 H2N2-imprinted individuals (95% CI: 4.14–5.20). This is significantly higher than the mean observed influenza mortality rates for 65 year-old H1N1_*β*_ and H1N1_*γ*_ cohorts in recent H1N1pdm09 seasons (1.52 deaths per 100,000; 95% CI: 1.75–1.29). As a counterfactual, if H1N1 never shifted into the 2009 pandemic virus, H3N2 and H2N2 cohorts would experience 0.15 (95% CI: 0.11–0.23) and 0.07 (95% CI: 0.05–0.11) deaths per 100,000, respectively, at the same age. Our results suggest that, as H3N2 and H2N2 cohorts age, and H1N1pdm09 continues to co-circulate with H3N2, we are likely to see substantially higher seasonal influenza mortality burden among older adults. H2N2- and H3N2-imprinted cohorts will continue to face higher mortality risk later in life compared to H1N1-imprinted cohorts, who have benefited from immune imprinting protection in old age.

## Discussion

Our results based on population-level influenza mortality data support the theory that immune imprinting and viral exposure history have long-standing impacts on influenza mortality risk over a cohort’s lifecourse. We take advantage of a long timeseries of single-year single-age mortality data to provide new insight into the effects of antigenic similarity on immune protection in the context of immune imprinting. When we decompose the mortality data by age and period, we find that the age profile of mortality rates closely follow observed influenza mortality rates prior to 2009, but observations deviate substantially from fitted rates for H1N1pdm09-dominant seasons. By focusing on mortality by single-year birth cohort rather than mortality by age group, we are able to identify transitions between positive and negative residuals during H1N1pdm09-dominant influenza seasons that are dependent on birth year, rather than age. The absence of similar cohort-period effects in the model fit to all-cause mortality and pancreatic cancer mortality confirms that the observed trends are specific to influenza mortality, rather than a general cohort or period effect.

Our results show that the antigenic structure of a virus has substantial effects on the degree of protection from immune imprinting, and that imprinting protection against seasonal strains is narrower – strain-level rather than group-level – than what has been observed for novel avian strains (*3, 5, 6*). Our analysis shows that individuals who were likely first exposed to different H1N1 variants circulating during the early twentieth century exhibit different mortality patterns during H1N1pdm09 influenza seasons. The H1N1_*⍺*_-imprinted cohorts, including those born during and directly after the 1918 pandemic, in particular, demonstrate the greatest levels of immune protection against the antigenically similar H1N1pdm09 virus, made evident by lower observed mortality rates, large residuals in the Lee-Carter model, and significantly lower age-adjusted mortality rates predicted by the negative binomial model. Cohorts more likely to have first encountered the later H1N1_*β*_ and H1N1_*γ*_ variants have relatively less protection against the 2009 pandemic virus, as seen by comparatively lower reductions in death counts and negative residuals in the Lee-Carter model during H1N1pdm09 seasons. This declining effect for cohorts first exposed to later H1N1 lineages suggests that viral evolution erodes the protective effects of immune imprinting against influenza mortality. Our observation echoes traditional effects of pathogen evolution on transmission-blocking immunity. Closer antigenic similarity between the virus first encountered in childhood and a virus encountered later in life provides advanced immune protection compared to more distant variants of influenza. Further, our results also provide support for antigenic seniority: repeated exposure to H1N1_*⍺*_ reinforces adaptive immune response, providing better protection to individuals who were exposed to early twentieth century H1N1 strains. Importantly, the lack of significant deviations in mortality during H3N2-dominant seasons, Influenza B-dominant seasons, and seasons with no dominant strain reinforces that our findings are specific to H1N1 imprinting.

The relatively modest protection experienced by H3N2-imprinted cohorts during H3N2-dominated seasons is likely due to the higher rate of antigenic evolution that has been observed for H3N2 viruses compared to other IAV strains (*28–30*). Our results also show that H3N2-imprinted cohorts experience relatively higher influenza mortality risk in seasons dominated by other strains. Consequently, compared to older H1N1- and H2N2-imprinted cohorts, H3N2 cohorts have experienced substantially higher age-specific influenza mortality through much of their life, and may continue to do so as long as H1N1pdm09 continues to circulate. This is a striking contrast to typical patterns in population health where younger cohorts generally experience lower mortality at all ages compared to older cohorts as life expectancy increases over time. These results also suggest that total mortality due to seasonal influenza is likely to increase as H2N2 and H3N2 cohorts age into an influenza landscape dominated by H3N2 and H1N1pdm09. Our results indicate that the H1N1pdm09 virus causes statistically higher flu-related mortality compared to pre-2009 H1N1 strains. The virulence of H1N1pdm09 may currently be masked as older cohorts, who are most likely to experience severe influenza morbidity and mortality, have immune protection against the virus. Finally, if avian H5N1 influenza viruses that are currently circulating widely in cattle and other mammals are able to adapt and cause human transmission, H3N2 cohorts may experience even greater mortality risk compared to older H1N1-imprinted cohorts (*3, 55–57*).

Our results also have implications for the impact of influenza vaccination. The observed cohort effects suggest that current influenza vaccines, which are updated seasonally in accordance with circulating strains, may not entirely decrease the elevated risk experienced by certain birth cohorts. While the observed effect of influenza imprinting on immune protection is strong, it provides narrow protection. Cohorts imprinted with a strain that is recirculating are not necessarily protected if the circulating strain does not have antigenic similarity to the imprinted virus.

With the expansion of influenza vaccination, many infants in the U.S. now receive an influenza vaccine before they experience an infection. Our results also underline the importance of future empirical studies to determine whether immune imprinting after vaccination (before any infection) is equivalent to that following from one’s first infection (*58*). If this is the case, and in the absence of a universal influenza vaccine (UIV) (see *e.g.* (*59–61*)), our results suggest that vaccinating naive children in such a way that ensures strong H1N1 response, while maintaining protection against other seasonal strains, may provide broader protection throughout their lives. More importantly, our results highlight the importance of the development and mass deployment of a UIV. In particular, a UIV that provides broad immune protection across influenza subtypes could lessen the mortality differentials observed between birth cohorts.

Our work also highlights important, yet previously unexplored, avenues for applying formal demographic methods in the study of infectious disease dynamics. To our knowledge, this study is the first to integrate a Lee-Carter model into the analysis of infectious disease mortality. Formal demographic methods allow us to decompose age and period effects and to analyze temporal population-level patterns in mortality. By modeling influenza mortality differentials at the population level, cohort-period effects that support the phenomenon of immune imprinting are incredibly apparent. Further implementation of Lee-Carter models and other demographic modeling methods could be highly beneficial in illustrating birth-year effects for other infectious diseases. Lee-Carter models may also help us study how imprinting to the neuraminidase of H1N1 affects morbidity and mortality to H5N1, and how antigenic drift of H1N1 affects NA protection relative to HA protection.

This study has several limitations that can be addressed in future work. First, we assign H1N1 lineage imprinting to cohorts spanning 1918–1953 by assuming that cohorts born within the circulation period of a specific H1N1 lineage were imprinted exclusively by that variant. This overlooks several important influenza transmission dynamics that may determine imprinting, such as individual risk factors, age of first exposure, regional variations, and co-circulation of antigenically-distinct strains (*3, 62*). Our implementation of the Gostic *et al.* (2016) (*3*) method to assign strain-specific imprinting is reliant on the accuracy of historical influenza surveillance data, and additionally assumes a constant risk of influenza exposure each season. Future research directions could benefit from correcting these simplifications, as more robust modeling of H1N1 lineage imprinting could reveal more subtle cohort-period effects in seasonal influenza mortality. Looking forward, our methods in conjunction with data from a potential Global Immunological Observatory (see (*63, 64*)) or from large cohort studies as suggested by (*65*), could clarify a number of these questions. Second, our Lee-Carter model estimates are based on historical data which have been dominated by H3N2 seasons. This means that the average age-profile is skewed towards the mortality observed during historical H3N2 outbreaks. However, this impact appears negligible, as evidenced by the minimal improvement in residual structure upon refitting the model separately to pre- and post-2009 data (Supplementary fig. S11). We are also able to explicitly account for this in our negative binomial regression framework, by including terms that capture how mortality varies by both circulating strain type and time period. An additional limitation of our Lee-Carter framework is that our bootstrap method resampled single-age and single-season death probabilities, rather than seasons, to preserve the ordered timeseries. This may underestimate parameter uncertainty by not fully capturing within-season correlations. Next, our reliance on viral specimen test data to determine the dominant seasonal strain is prone to biases as strains that cause less severe morbidity may be under-tested for, artificially raising the prevalence of more severe strains. Finally, a major limitation of our work is the reliance on mortality data to estimate relative risk between cohorts. The availability of a long time series of mortality data for the entire population by single-year age groups allows us to precisely identify birth-year effects. However, mortality data capture only the most severe cases, and these results cannot be generalized to estimate relative risk for milder cases, which account for the vast majority of influenza burden. Mortality data also do not allow us to consider other factors that affect influenza incidence and mortality, such as co-morbidities, contact patterns, and health-seeking behavior, which may differ by population subgroups. ICD-coded mortality data are also limited by possible misclassifications of other respiratory diseases, and variations in coding practices. Expanded influenza surveillance efforts, along with the routine collection of serological data, will be crucial for understanding population-level risk patterns for influenza infection.

The findings of this population-level study on cohort imprinting effects on influenza mortality indicate that antigenic similarity between the strain an individual is imprinted with and the dominant strain circulating in an influenza season has a substantial effect on mortality risk. We show large differences in a cohort’s influenza mortality risk over their lifetime based on the interactions between immune imprinting, age-specific risk, and the circulating strain. Applying formal demographic tools to population-level data provides a unique opportunity to disentangle biological mechanisms that determine risk of mortality from infectious diseases, and to understand how age, period, and cohort effects shape a population’s mortality patterns.

## Data Availability

All data produced are publicly available online from the National Center for Health statistics (https://www.cdc.gov/nchs/data_access/vitalstatsonline.htm), Human Mortality Database (https://www.mortality.org), and FluNet (https://www.who.int/tools/flunet)
All code for the analysis is available on Github at: \\ \url{https://github.com/kylee-hoffman/influenza-imprinting-public}.

## Funding

A.S.M. and K.A.H. received support for this research from the National Institute of General Medical Sciences of the National Institutes of Health under award number R35GM156856. A.S.M. and K.A.H. received support from the National Institute of Aging of the National Institutes of Health under award number R01AG058940. C.M.S.-R. gratefully acknowledges funding from the Miller Institute for Basic Research in Science of UC Berkeley via a Miller Research Fellowship. K.A.H. received support from the National Institute for Child Health and Human Development of the National Institutes of Health under award numbers 5T32HD101364 and T32-HD007275.

## Author contributions

K.A.H. and A.S.M. designed the study. K.A.H. conducted the analyses. C.M.S.-R. assisted with the methodology and conceptualization. A.S.M. assisted with the analysis and oversaw the study. All authors wrote and edited the manuscript.

## Competing interests

There are no competing interests to declare.

## Data and materials availability

All data are publicly available from the National Center for Health statistics (mortality), Human Mortality Database (exposure), and FluNet (strain dominance), and can be downloaded from:

https://www.cdc.gov/nchs/data_access/vitalstatsonline.htm,

https://www.mortality.org,

https://www.who.int/tools/flunet.

All code for the analysis is available on Github at:

https://github.com/kylee-hoffman/influenza-imprinting-public.

## Notes

### Competing Interest Statement

The authors have declared no competing interest.

### Author Declarations

The study used ONLY openly available human data that were originally located at: https://www.cdc.gov/nchs/data_access/vitalstatsonline.htm#Mortality_Multiple

