## Supplementary Material for "Childhood immune imprinting shapes cohort and period influenza mortality"

**This PDF file includes:**

Figures S1 to S12

Tables S1 to S4

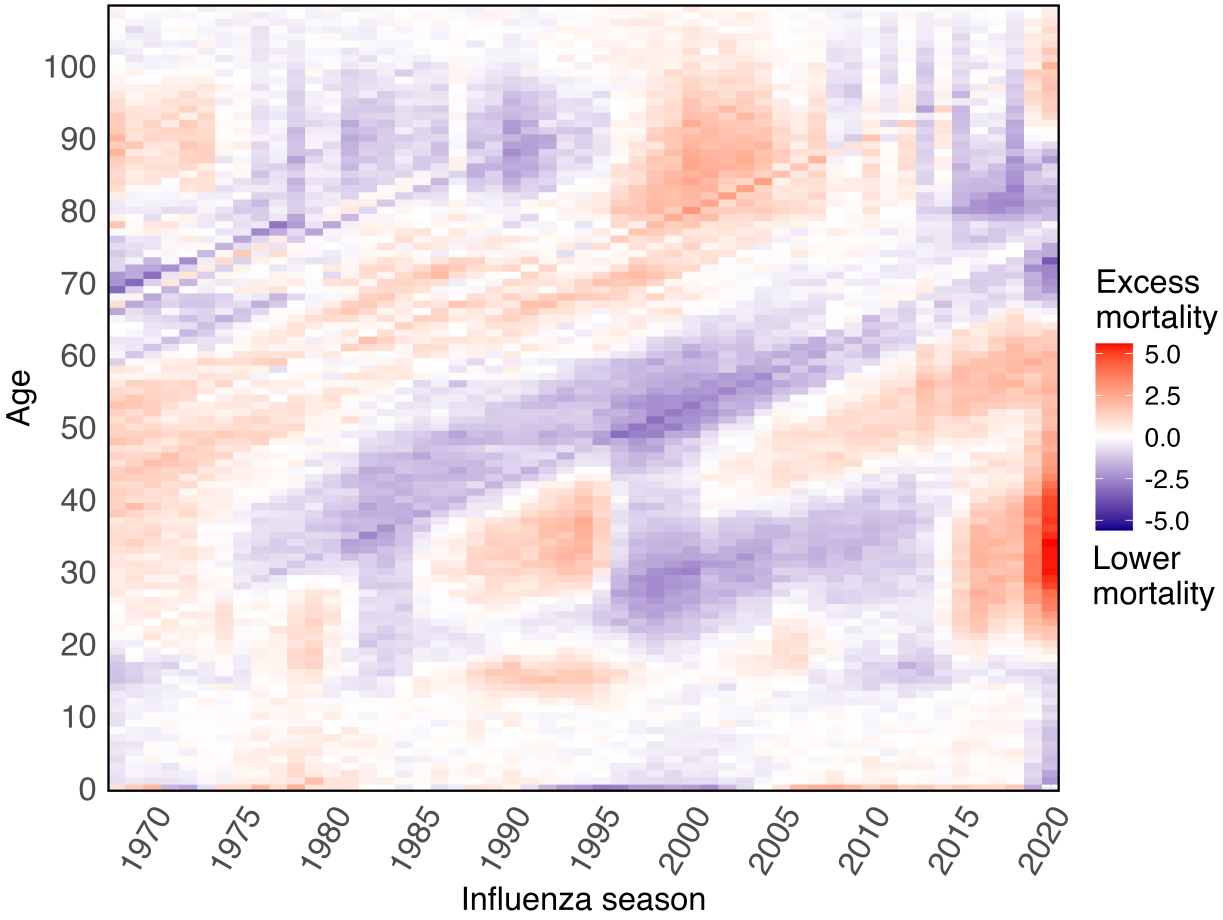


**Figure S1**: **Deviance residuals of the Lee-Carter model for all-cause mortality by age and season.** Positive residuals (in red) indicate higher observed age-specific mortality rates in comparison to the Lee-Carter estimated rates; negative residuals (in blue) indicate lower observed mortality than estimated. Diagonal banding indicate birth cohort effects in all cause mortality.

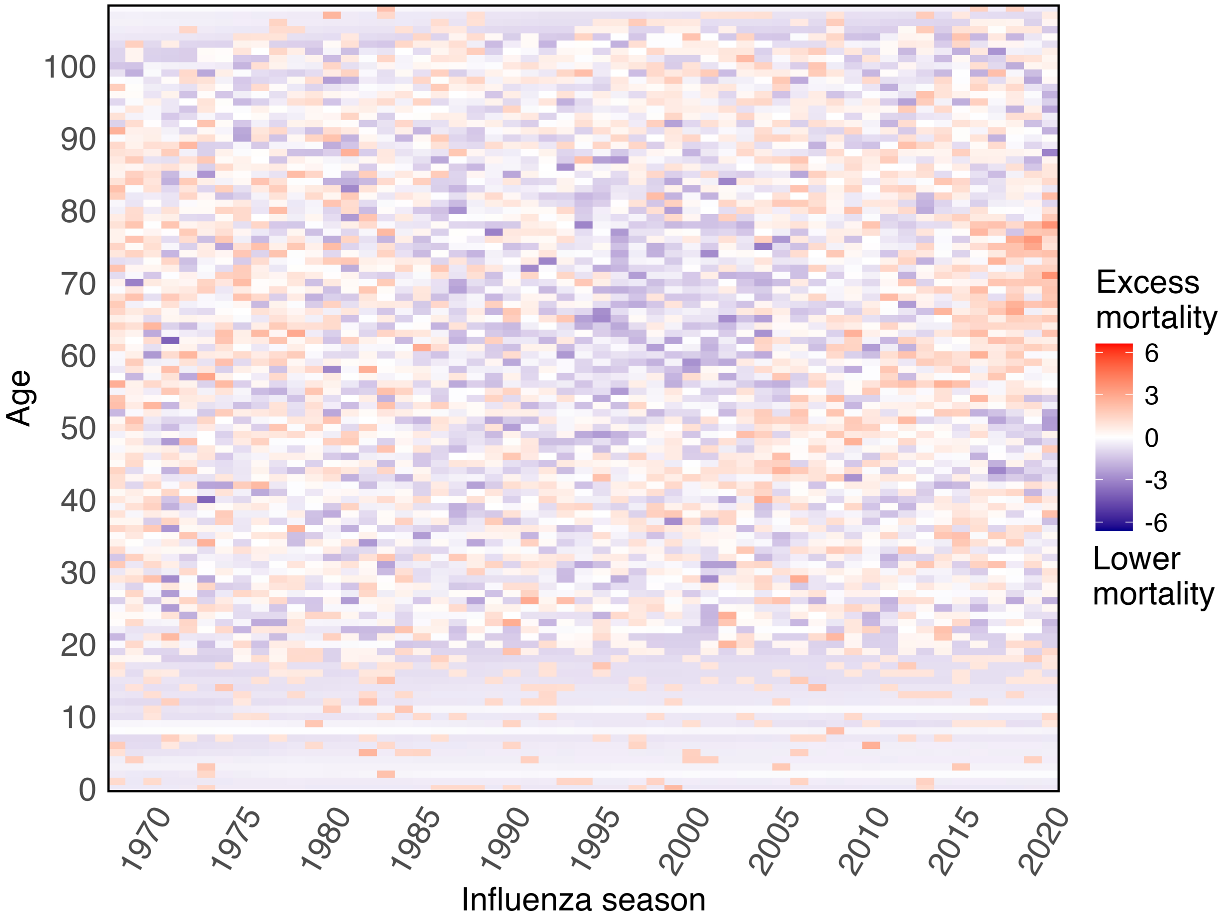


**Figure S2**: **Deviance residuals of the Lee-Carter model for pancreatic cancer mortality by age and season.** Same as Figure S1, but for pancreatic cancer mortality.

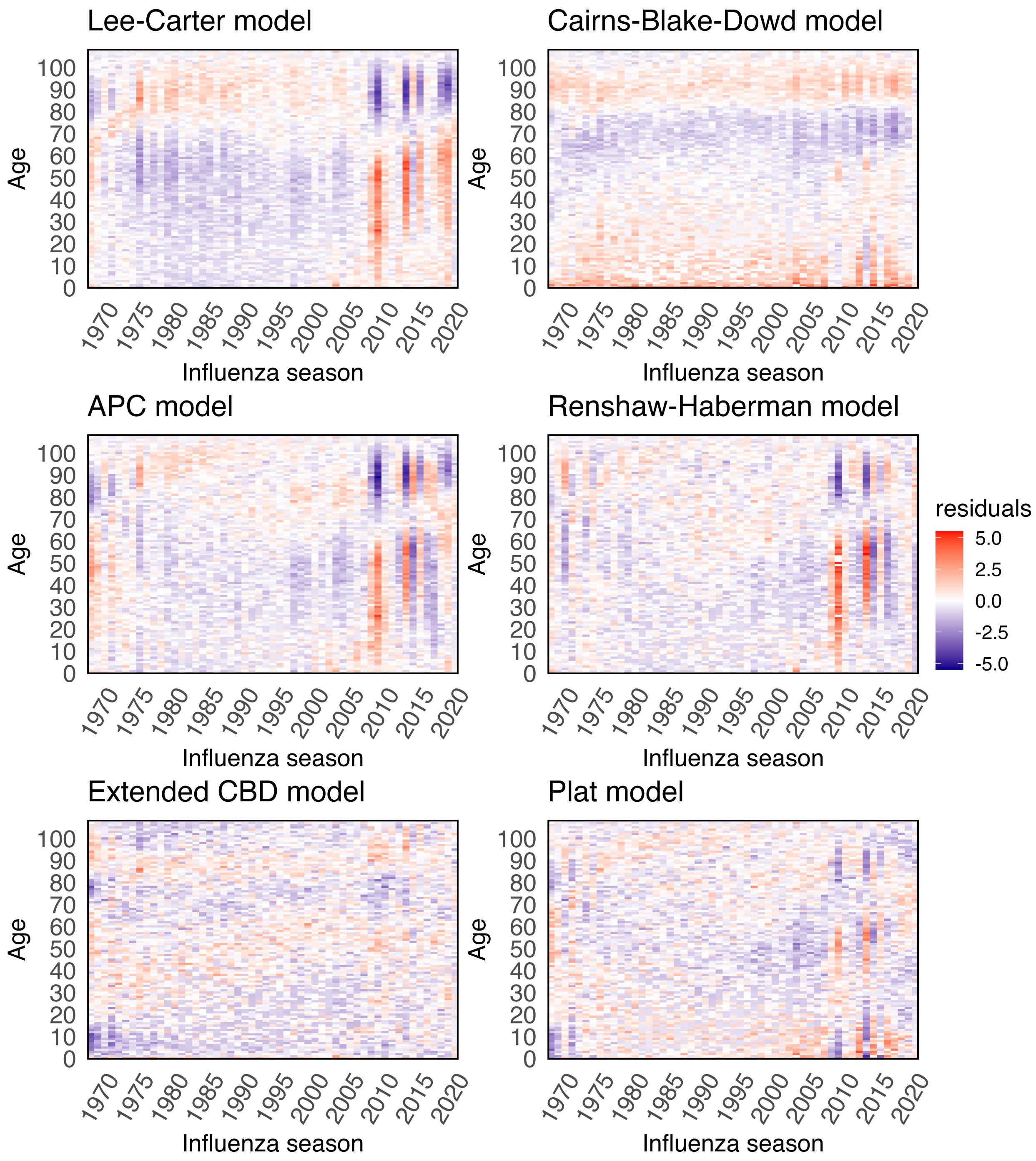


**Figure S3**: **Deviance residuals for influenza mortality from different stochastic mortality models.** Same as Figure S1, but for different models. All models except Lee-Carter and Cairns-Blake-Dowd include a cohort term.

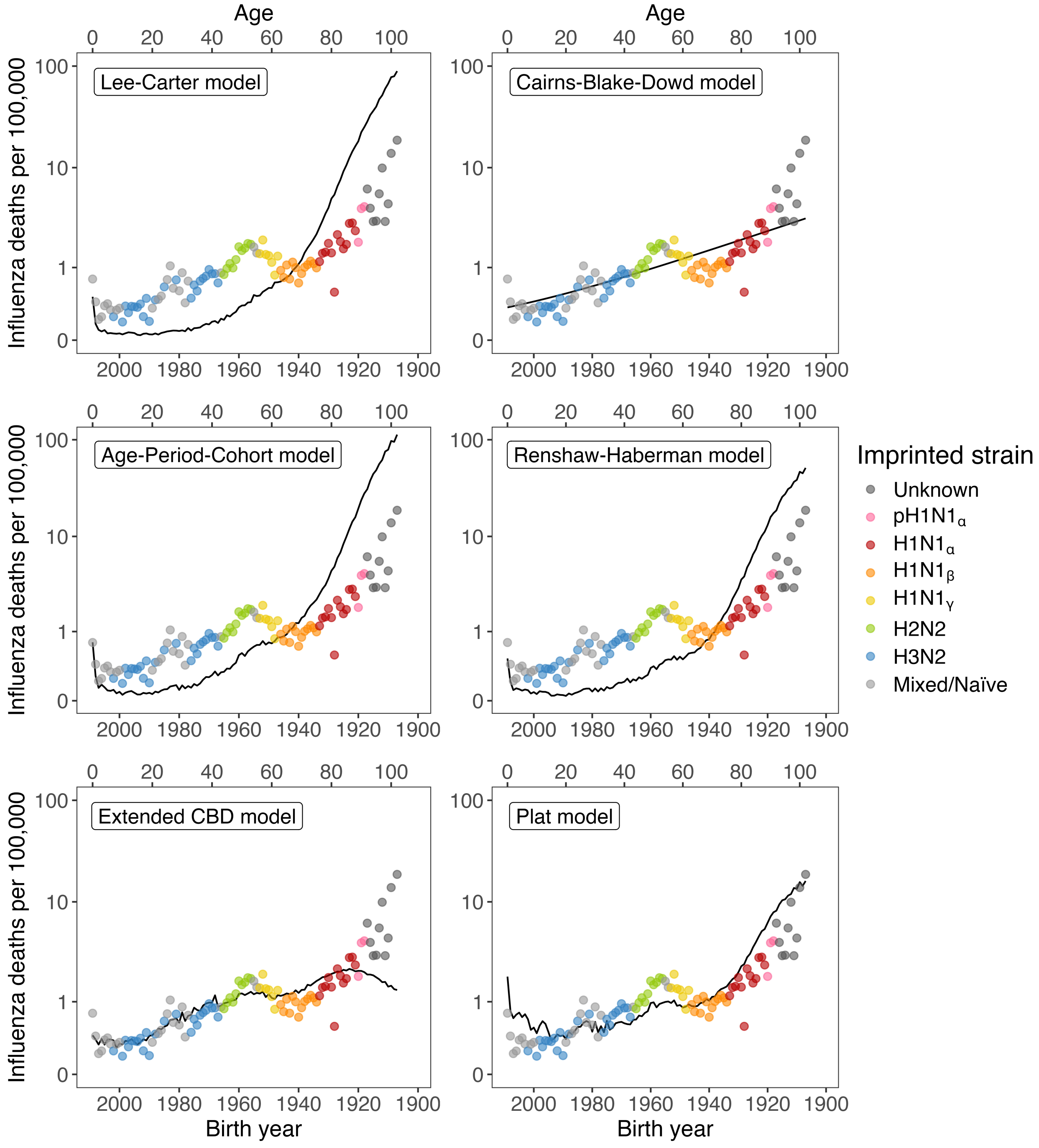


**Figure S4**: **Stochastic mortality model fits to 2009-2010 in-sample data.** All models except Lee-Carter and Cairns-Blake-Dowd include a cohort term. The APC model and Renshaw-Haberman model fitted rates are visually similar to the Lee-Carter modeled rates. The Plat model fits the 2009-2010 season exceptionally well. This could be due to constrains of the Plat model preventing the cohort term from compensating for missing age-period effects.

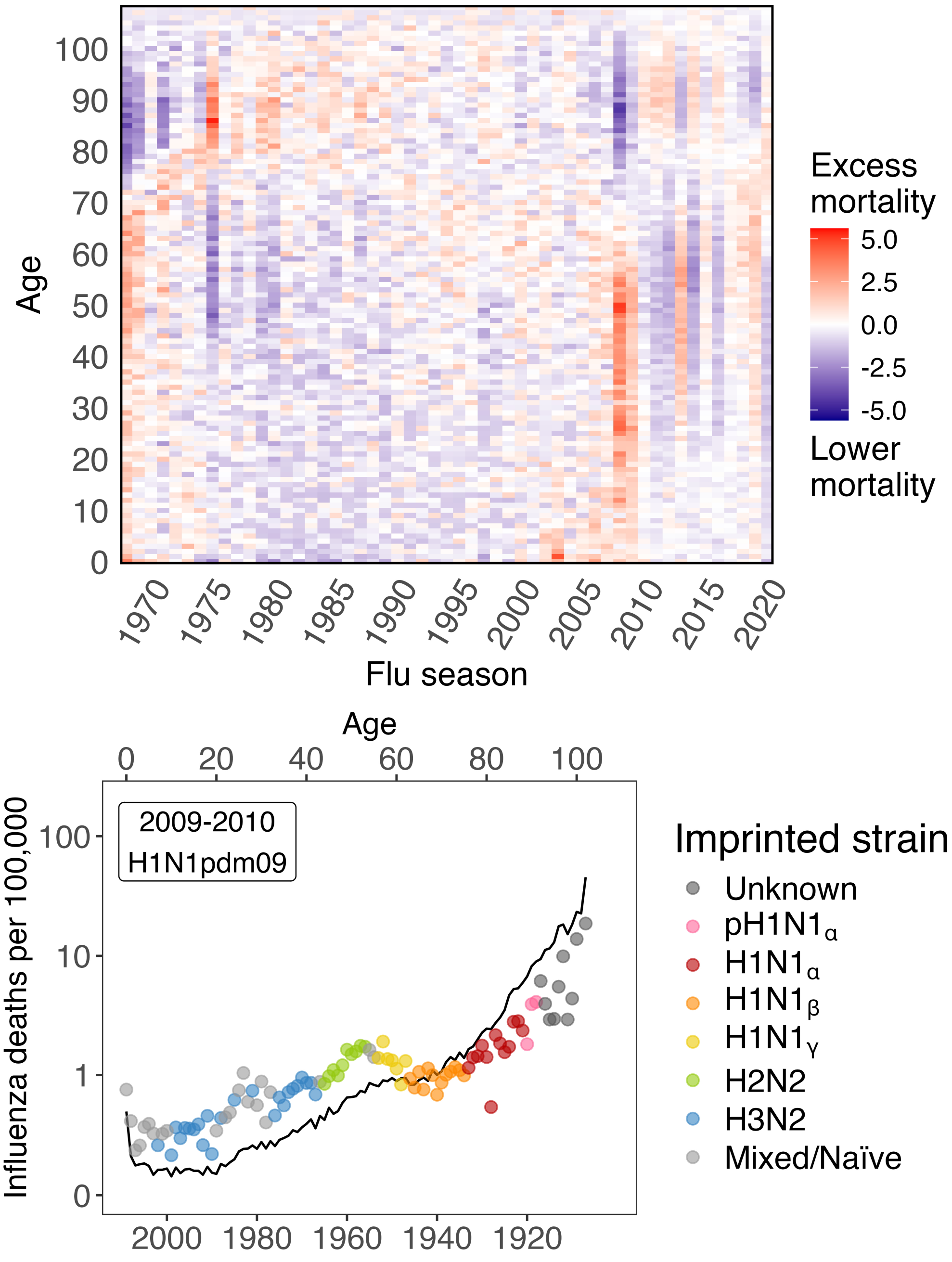


**Figure S5**: **Deviance residuals of two Lee-Carter models fitted separately to pre- and post-2009 influenza mortality.** The heat map is the same as Figure S1, but stitches together the residuals from both models: 1968-2008 for the first model, and 2009-2020 for the second model. The bottom-most plot shows observed age-specific mortality rates for the 2009-2010 influenza season compared to fitted rates produced by the model only fit to data from 2009 and beyond.

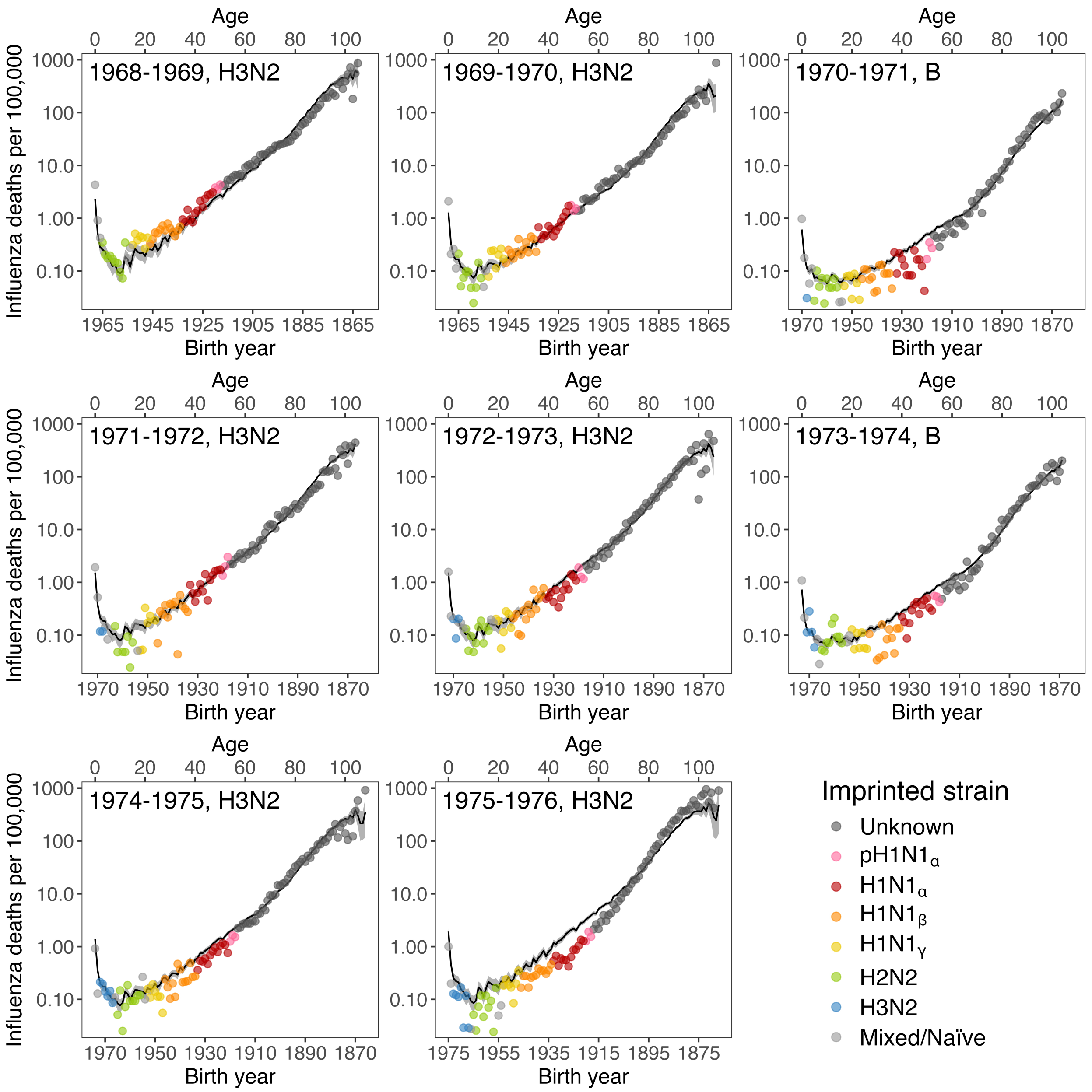


**Figure S6**: **Lee-Carter estimated influenza mortality rates per 100,000 population compared to observed rates for all individual influenza seasons between 1968 and 1976.** Solid black line represents the expected rates with 95 percent confidence intervals based on the Lee-Carter model fit, and each point represents the observed single-year age-specific mortality rate for a birth cohort.

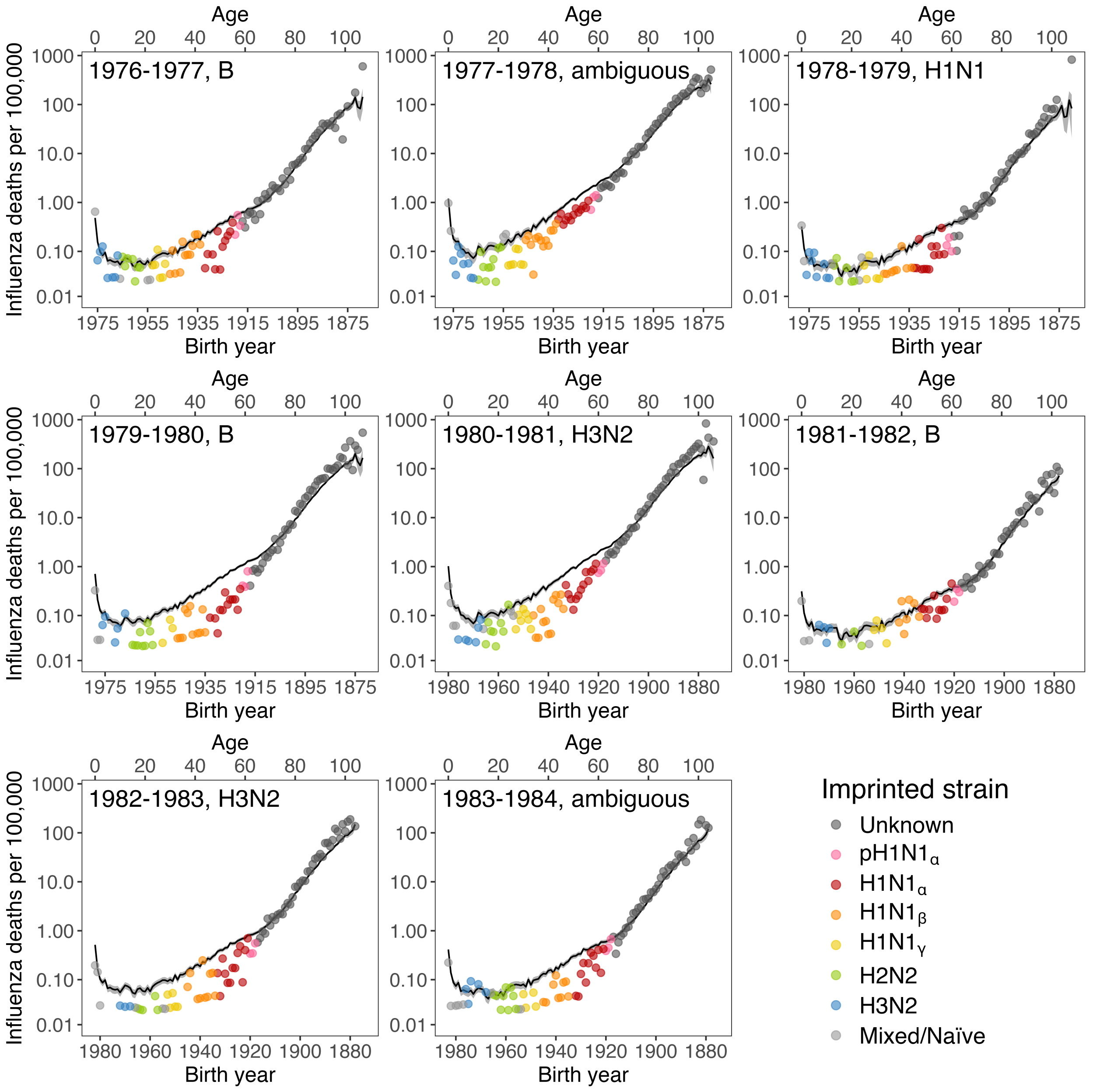


**Figure S7**: **Lee-Carter estimated influenza mortality rates per 100,000 population compared to observed rates for all individual influenza seasons between 1976 and 1984.** Same as Figure S6, but for seasons between 1976 and 1984.

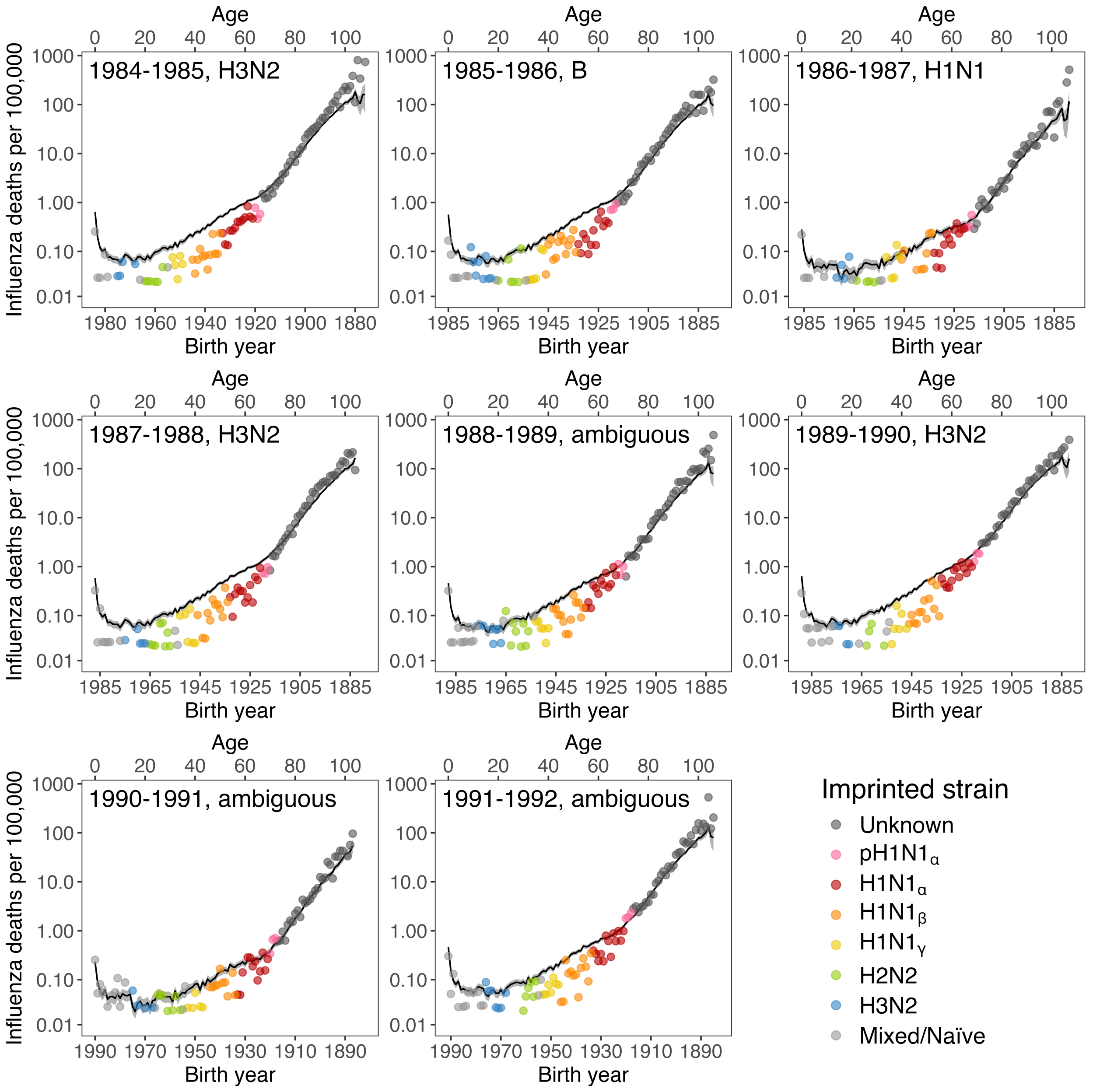


**Figure S8**: **Lee-Carter estimated influenza mortality rates per 100,000 population compared to observed rates for all individual influenza seasons between 1984 and 1992.** Same as Figure S6, but for seasons between 1984 and 1992.

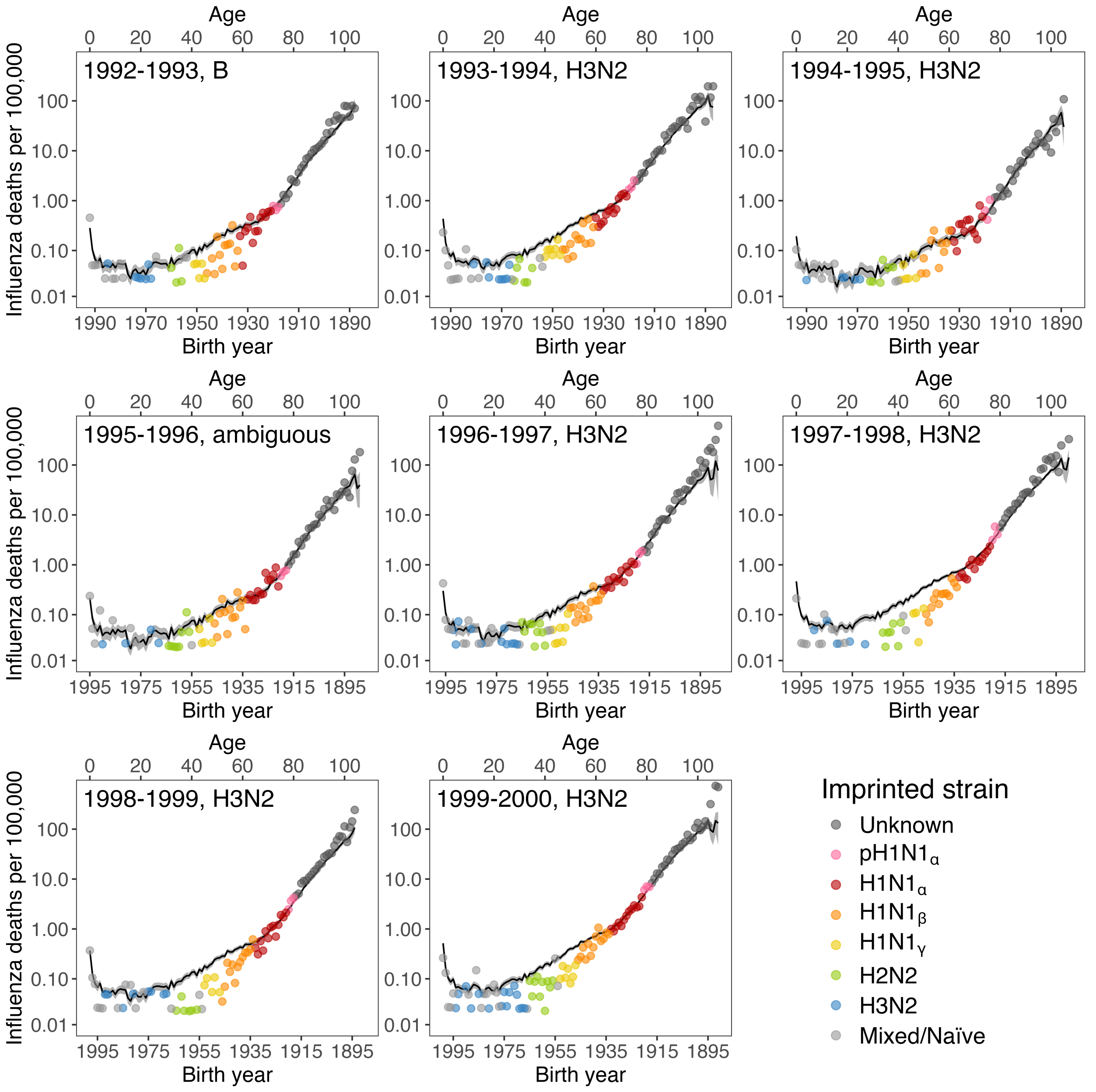


**Figure S9**: **Lee-Carter estimated influenza mortality rates per 100,000 population compared to observed rates for all individual influenza seasons between 1992 and 2000.** Same as Figure S6, but for seasons between 1992 and 2000.

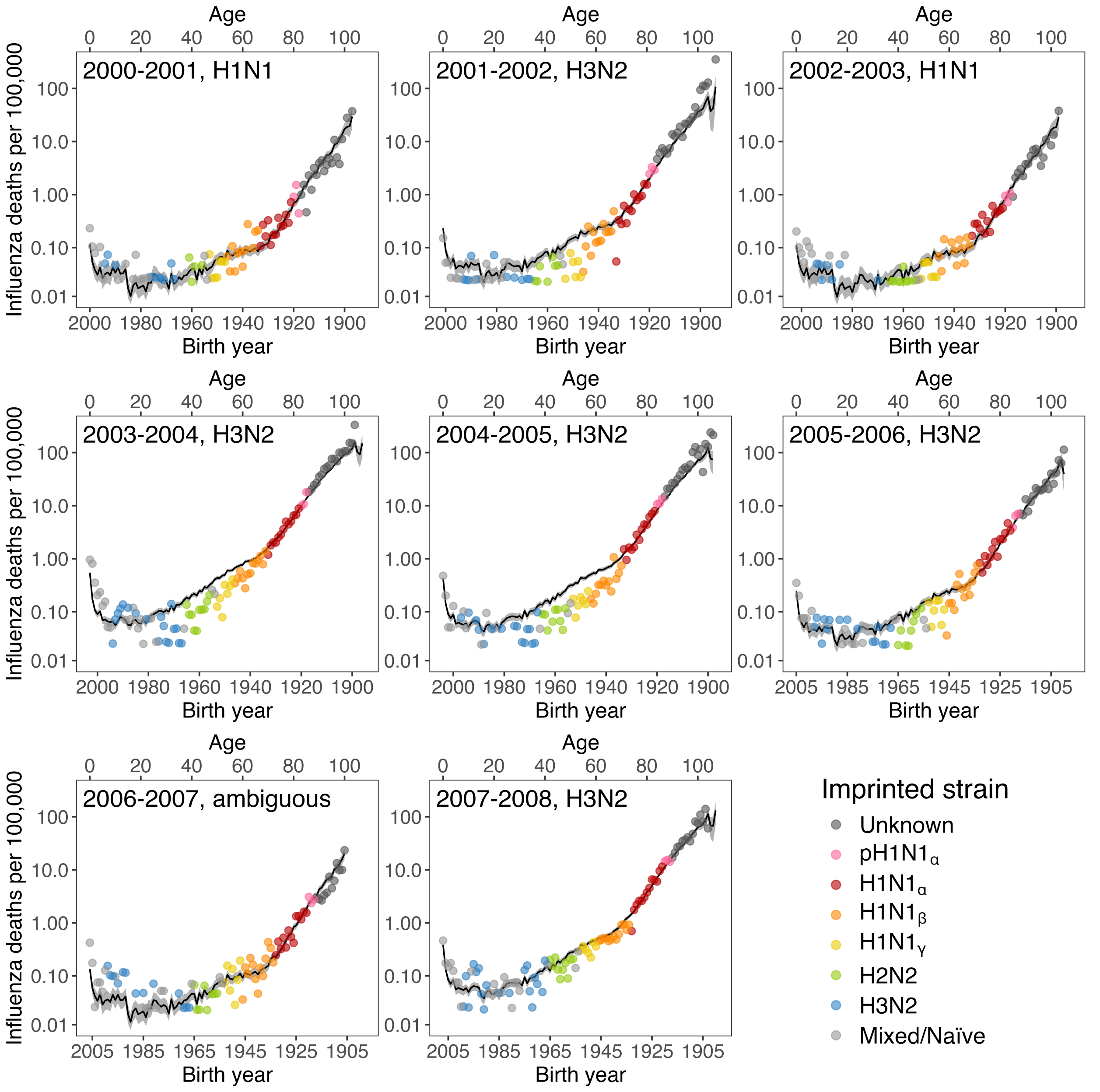


**Figure S10**: **Lee-Carter estimated influenza mortality rates per 100,000 population compared to observed rates for all individual influenza seasons between 2000 and 2008.** Same as Figure S6, but for seasons between 2000 and 2008.

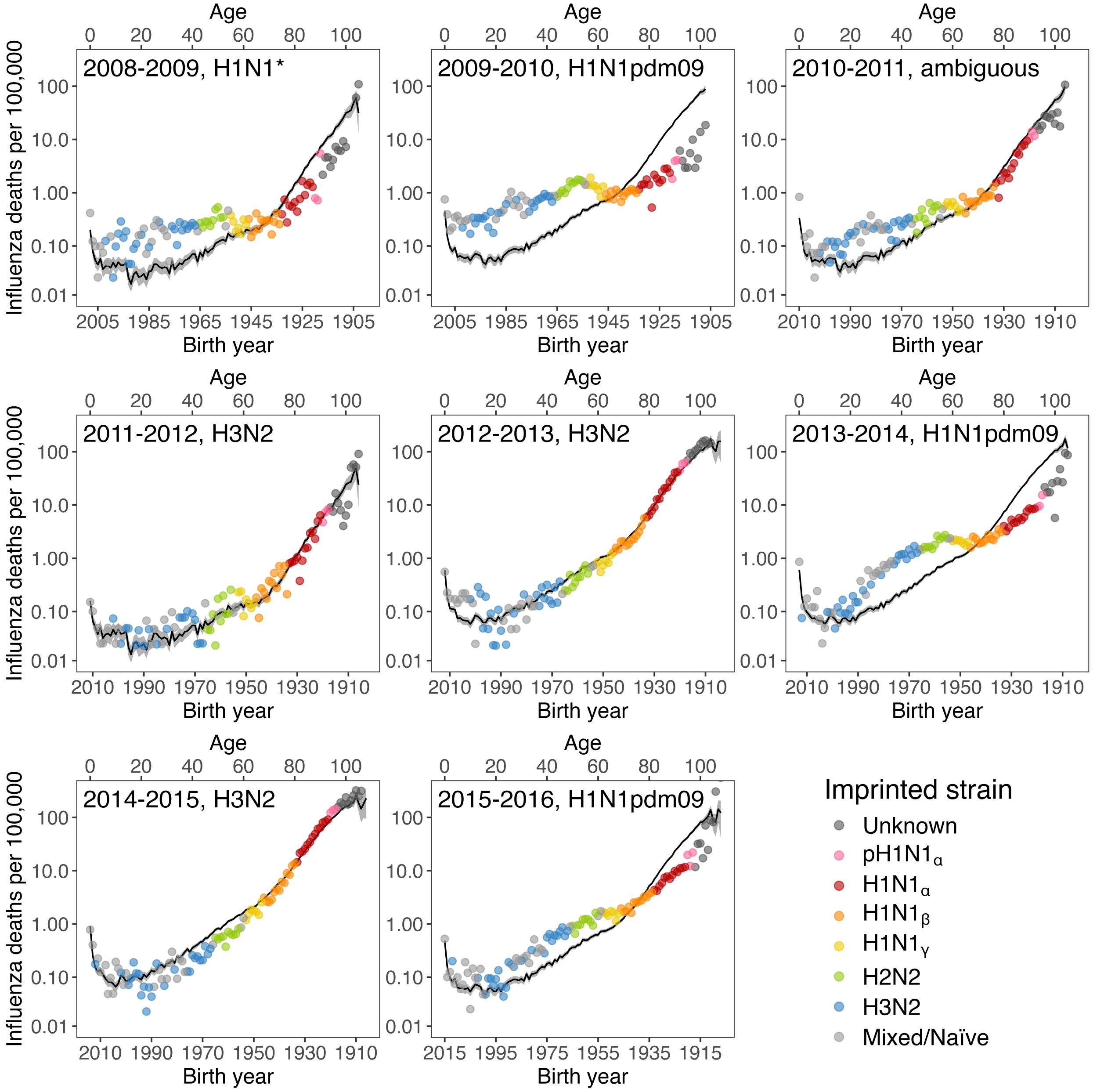


**Figure S11**: **Lee-Carter estimated influenza mortality rates per 100,000 population compared to observed rates for all individual influenza seasons between 2008 and 2016.** Same as Figure S6, but for seasons between 2008 and 2016.

*Seasonal H1N1 was the dominant strain for the 2008-2009 influenza season until April 2009, when H1N1pdm09 was first detected and overtook previous H1N1 strains.

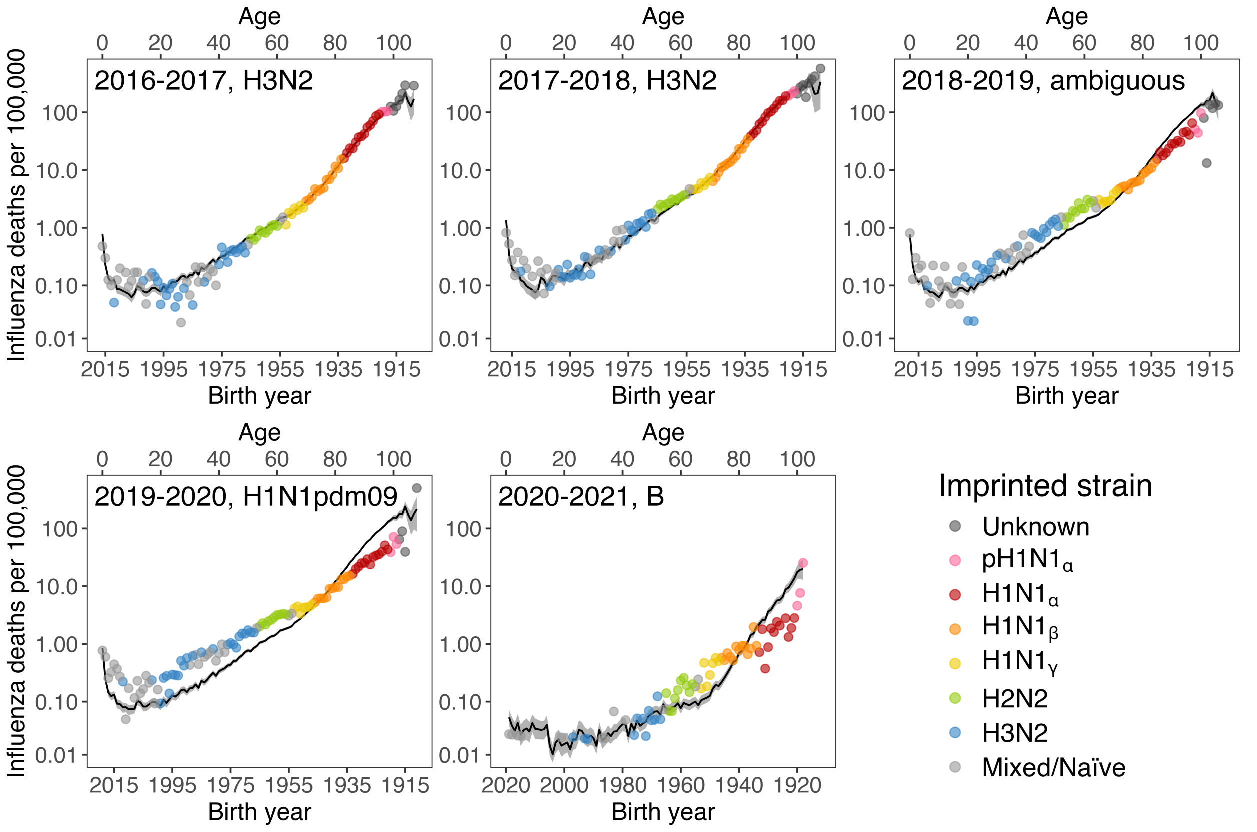


**Figure S12**: **Lee-Carter estimated influenza mortality rates per 100,000 population compared to observed rates for all individual influenza seasons between 2016 and 2021.** Same as Figure S6, but for seasons between 2016 and 2021.

**Table S1**: **Rate ratios (RR), robust standard errors and 95% confidence intervals estimated from a negative binomial regression model.** The model also included a natural spline for ages and a categorical variable for season.

| *Independent variable* | | Rate ratio | Robust SE | 95% CI |
|---|---|---|---|---|
| Imprinted strain | Circulating strain | | | |
| H3N2 | - | Reference | - | - |
| pH1N1$_\alpha$ | - | 1.088 | 0.097 | 0.913–1.297 |
| H1N1$_\alpha$ | - | 0.941 | 0.073 | 0.808–1.097 |
| H1N1$_\beta$ | - | 0.807 | 0.055 | 0.706–0.923 |
| H1N1$_\gamma$ | - | 0.807 | 0.051 | 0.712-0.914 |
| H2N2 | - | 0.780 | 0.046 | 0.695–0.874 |
| Mixed | - | 1.085 | 0.055 | 0.981–1.199 |
| Unknown | - | 1.398 | 0.124 | 1.175–1.663 |
| - | H3N2 | Reference | - | - |
| - | H1N1 | 0.031 | 0.006 | 0.022–0.052 |
| - | H1N1pdm09 | 1.502 | 0.428 | 0.860–2.625 |
| - | Ambiguous | 0.464 | 0.047 | 0.380–0.567 |
| pH1N1$_\alpha$ | H1N1 | 1.191 | 0.468 | 0.552–2.571 |
| pH1N1$_\alpha$ | H1N1pdm09 | 0.119 | 0.052 | 0.050–0.279 |
| pH1N1$_\alpha$ | Ambiguous | 0.438 | 0.058 | 0.338–0.567 |
| H1N1$_\alpha$ | H1N1 | 1.173 | 0.402 | 0.599–2.295 |
| H1N1$_\alpha$ | H1N1pdm09 | 0.179 | 0.061 | 0.092–0.347 |
| H1N1$_\alpha$ | Ambiguous | 0.409 | 0.042 | 0.335–0.499 |
| H1N1$_\beta$ | H1N1 | 1.083 | 0.313 | 0.615–1.908 |
| H1N1$_\beta$ | H1N1pdm09 | 0.359 | 0.073 | 0.241–0.535 |
| H1N1$_\beta$ | Ambiguous | 0.504 | 0.049 | 0.417–0.609 |
| H1N1$_\gamma$ | H1N1 | 1.003 | 0.244 | 0.623–1.617 |
| H1N1$_\gamma$ | H1N1pdm09 | 0.586 | 0.055 | 0.488–0.705 |
| H1N1$_\gamma$ | Ambiguous | 0.666 | 0.073 | 0.538–0.824 |
| H2N2 | H1N1 | 0.607 | 0.147 | 0.377–0.976 |
| H2N2 | H1N1pdm09 | 0.920 | 0.095 | 0.751–1.128 |
| H2N2 | Ambiguous | 1.013 | 0.109 | 0.820–1.251 |
| Cumulative exposure $\times$ Circulating strain | | | | |
| - | H3N2 | 0.554 | 0.117 | 0.366–0.838 |
| - | H1N1 | 0.240 | 0.161 | 0.065–0.052 |
| - | H1N1pdm09 | 0.123 | 0.167 | 0.009–1.763 |
| - | Ambiguous | - | - | - |

**Table S2**: **Circulating strain dominance for influenza seasons between 1968-2021.** Compiled from CDC Influenza Surveillance Report archives, available through CDC Stacks (1968–1996), and WHO FluNet data portal (1996–2021).
*Seasonal H1N1 was the dominant strain for the 2008-2009 influenza season until April 2009, when H1N1pdm09 was first detected and overtook previous H1N1 strains.

| Influenza season | Dominant circulating strain | Influenza season | Dominant circulating strain |
|---|---|---|---|
| 1968–1969 | H3N2 | 1996–1997 | H3N2 |
| 1969–1970 | H3N2 | 1997–1998 | H3N2 |
| 1970–1971 | B | 1998–1999 | H3N2 |
| 1971–1972 | H3N2 | 1999–2000 | H3N2 |
| 1972–1973 | H3N2 | 2000–2001 | H1N1 |
| 1973–1974 | B | 2001–2002 | H3N2 |
| 1974–1975 | H3N2 | 2002–2003 | H1N1 |
| 1975–1976 | H3N2 | 2003–2004 | H3N2 |
| 1976–1977 | B | 2004–2005 | H3N2 |
| 1977–1978 | Ambiguous | 2005–2006 | H3N2 |
| 1978–1979 | H1N1 | 2006–2007 | Ambiguous |
| 1979–1980 | B | 2007–2008 | H3N2 |
| 1980–1981 | H3N2 | 2008–2009 | H1N1* |
| 1981–1982 | B | 2009–2010 | H1N1pdm09 |
| 1982–1983 | H3N2 | 2010–2011 | Ambiguous |
| 1983–1984 | Ambiguous | 2011–2012 | H3N2 |
| 1984–1985 | H3N2 | 2012–2013 | H3N2 |
| 1985–1986 | B | 2013–2014 | H1N1pdm09 |
| 1986–1987 | H1N1 | 2014–2015 | H3N2 |
| 1987–1988 | H3N2 | 2015–2016 | H1N1pdm09 |
| 1988–1989 | Ambiguous | 2016–2017 | H3N2 |
| 1989–1990 | H3N2 | 2017–2018 | H3N2 |
| 1990–1991 | Ambiguous | 2018–2019 | Ambiguous |
| 1991–1992 | Ambiguous | 2019–2020 | H1N1pdm09 |
| 1992–1993 | B | 2020–2021 | B |
| 1993–1994 | H3N2 | | |
| 1994–1995 | H3N2 | | |
| 1995–1996 | Ambiguous | | |

**Table S3**: **Influenza A variant imprinting by birth year**. Birth cohorts born within or shortly before the study period are typically classified as immunologically naïve to IAV in their first years of childhood. This table provides the most probable imprinted strain after naïveté.

| Variant | Birth years | N |
|---|---|---|
| Unknown | 1860-1917 | 58 |
| pH1N1$_\alpha$ | 1918-1920 | 3 |
| H1N1$_\alpha$ | 1921-1933 | 13 |
| H1N1$_\beta$ | 1934-1946 | 13 |
| H1N1$_\gamma$ | 1947-1953 | 7 |
| H2N2 | 1956-1965 | 10 |
| H3N2 | 1967-1976, 1981, 1985, 1988, 1990-1999, 2002, 2012 | 25 |
| Mixed | 1954-1955, 1966, 1977-1980, 1982-1984, 1986-1987, 1989, 2000-2001, 2003-2008, 2010-2011, 2013-2015 | 28 |

**Table S4**: **Stochastic mortality model structures and performance**. Akaike information criteria based on fitting each model to all seasons between 1968-1969 and 2020-2021. Parameters not described elsewhere include: $\gamma_{t-x}$: cohort term; $\bar{x}$: average age in data; $\hat{\sigma}_x^2$: average value of $(x-\bar{x})^2$. Each model was originally formulated to estimate and forecast all-cause mortality. All models were fit using the R package StMoMo (*46*).

| Model | Formula | ΔAIC |
|---|---|---|
| Extended CBD | $\text{logit}(q_{x,t}) = k_t^{(1)} + (x-\bar{x})k_t^{(2)} + [(x-\bar{x})^2 - \hat{\sigma}_x^2]k_t^3 + \gamma_{t-x}$ | 0.0 |
| Plat | $\text{logit}(q_{x,t}) = a_x + k_t^{(1)} + (\bar{x}-x)k_t^{(2)} + \gamma_{t-x}$ | 1394.0 |
| Renshaw-Haberman | $\text{logit}(q_{x,t}) = a_x + b_x^{(1)}k_t + \gamma_{t-x}$ | 7765.4 |
| APC | $\text{logit}(q_{x,t}) = a_x + k_t + \gamma_{t-x}$ | 12475.9 |
| Lee-Carter | $\text{logit}(q_{x,t}) = a_x + b_x k_t$ | 20398.7 |
| Cairns-Blake-Dowd | $\text{logit}(q_{x,t}) = k_t^{(1)} + (x-\bar{x})k_t^{(2)}$ | 24553.4 |
